# Targeting Multiple Conserved T-Cell Epitopes for Protection against COVID-19 Moderate-Severe Disease by a Pan-Sarbecovirus Vaccine

**DOI:** 10.1101/2023.06.28.23291948

**Authors:** Chang Yi Wang, Be-Sheng Kuo, Yu-Hsiang Lee, Yu-Hsin Ho, Yi-Hua Pan, Ya-Ting Yang, Hsi-Chi Chang, Lin-Fang Fu, Wen-Jiun Peng

**Affiliations:** UBI Asia, Hsinchu, Taiwan

**Author notes:** To whom correspondence should be addressed Chang Yi Wang, Ph.D. Chairperson, Chief Scientific Officer United Biomedical, Inc. Currently on assignment at UBI Asia based in Taiwan.

**Keywords:** Pan-Sarbecovirus, SARS-CoV-2, COVID-19, UB-612, booster vaccination, and conserved T cell epitopes

## Abstract

**Background:** Most of current approved vaccines, based on a Spike-only as single immunogen, fall short of producing a full-blown T-cell immunity. SARS-CoV-2 continues to evolve with ever-emergent higher-contagious mutants that may take a turn going beyond Omicron to bring about a new pandemic outbreak. New recombinant SARS-CoV-2 species could be man-made through genetic manipulation to infect systemically. Development of composition-innovated, pan-variant COVID-19 vaccines to prevent from hospitalization and severe disease, and to forestall the next pandemic catastrophe, is an urgent global objective.

**Methods and findings:** In a retrospective, e-questionnaire Observational Study, extended from a clinical Phase-2 trial conducted in Taiwan, during the prime time of Omicron outbreak dominated by BA.2 and BA.5 variants, we investigated the preventive effects against COVID-19 moderate-severe disease (hospitalization and ICU admission) by a pan-Sarbecovirus vaccine UB-612 that targets monomeric S1-RBD-focused subunit protein and five designer peptides comprising sequence-conserved, non-mutable helper and cytotoxic T lymphocyte (Th/CTL) epitopes derived from Spike (S2), Membrane (M) and Nucleocapsid (N) proteins. Per UB-612 vaccination, there were no hospitalization and ICU admission cases (0% rate, 6 months after Omicron outbreak) reported ≥14 months post-2^nd^ dose of primary series, and ≥10 months post-booster (3^rd^ dose), to which the potent memory cytotoxic CD8 T cell immunity may be the pivotal in control of the infection disease severity. Six months post-booster, the infection rate (asymptomatic and symptomatic mild) was only 1.2%, which increased to 27.8% observed ≥10 months post-booster. The notable protection effects are in good alignment with a preliminary Phase-3 heterologous booster trial report showing that UB-612 can serve as a competent booster substitute for other EUA-approved vaccine platforms to enhance their seroconversion rate and viral-neutralizing titer against Omicron BA.5.

**Conclusions:** UB-612, a universal multitope vaccine promoting full-blown T cell immunity, may work as a competent primer and booster for persons vulnerable to Sarbecovirus infection.

**Trial Registration:** ClinicalTrials.gov ID: NCT04773067.

**AUTHOR SUMMARY:** A COVID-19 vaccine based on a Spike-only single immunogen would fall short of producing a full-blown, escape-proof T cell immunity. In Omicron era plagued with ever-evolving and higher-contagious SARS-CoV-2 mutants, immune antibodies against variants beyond BA.5 are seen on a cliff drop, rendering the viral-neutralizing titer strength an increasingly less relevant immunity parameter. The true, urgent issue at heart in vaccine development has not been updating variant component to increase antibody titer for prevention of infection, but to validate universal vaccines that would have a potential to head off hospitalization, severe disease and ultimately reinfection altogether, and so to forestall a new catastrophe of pandemic outbreak. To reach the ideal goals, a universal vaccine able to produce potent, broadly recognizing and durable memory T cell immunity would be essential. UB-612, a pan-Sarbecovirus T cell immunity-promoting mutitope vaccine, has been shown to provide strong and long-lasting ≥10 month protective effect against COVID-19 moderate-severe disease (0% cases of hospitalization and ICU admission). UB-612 is a unique S1-RBD subunit protein vaccine armed with five designer peptides comprising sequence-conserved helper and cytotoxic T lymphocyte (Th/CTL) epitopes derived from Spike (S2×3), Membrane (M) and Nucleocapsid (N) proteins across Sarbecovirus species.

## INTRODUCTION

The once-dominant Omicron BA.2 and BA.5 infections have been phased out and are followed by the higher-contagious subvariants related to BQ.1 and XBB, yet the numbers of hospitalization and death are on the decline under immunity influence from vaccination and infection. The present lull in COVID-19 incidence is signaling a downward trend of the pandemic after a 3-year catastrophe, echoed by an end to global “health emergency,” as declared by WHO on May 5, 2023 [1]. Overall, current authorized COVID-19 vaccines could have helped reduce severe disease and deaths, with primary vaccination and booster dose reportedly that could lower hospitalization rates by >10 and >2 folds, respectively, as compared to un-vaccination and without booster doses [2].

Nonetheless, the “pandemic” in a broader sense is far from over as virus remains evolving (**S1A Fig**) and surging in a number of disparate parts of the world [3-7]. In fact, the antibody-evading Omicron BQ.1 and XBB related variants are still circulating even in highly vaccinated regions of the world (**S1B Fig**). Given the case number of infection subsides for now, the lasting legacies of pandemic excess mortality [8-11], augmented by long COVID and vaccine injuries, remain to be dealt with. The worsening reinfection, responsible for the large swath of the COVID complications and sequelae, is created by the pathologically milder Omicrons.

The Omicron XBB family, emerged from a recombination of two BA.2 lineages (BA.2.10.1 and BA.2.75) [12], is currently leading the low-profile trajectory of COVID-19, with the higher-contagious XBB.1.5 dominating the course of infection and XBB.1.16 gaining ground of new infection cases in USA (**S1C Fig**). Relative to the parent XBB, the XBB.1.16 acquires three additional mutations (E180V, F486P and K478R) that could account for a 1.2-fold higher antibody-evasion than XBB.1.5 [13,14]without significant between-Omicron differences in symptom scope and disease severity [15].

Relative to Delta strain, Omicron family presents a feeble attack to lung tissues and thus a lower pathogenicity, due to inefficient cell-to-cell syncytia after landing on upper respiratory membrane and differential cell entry mechanism without use of fusion co-receptor TMPRSS2 [16-18]. However, the Omicron COVID-19 is of non-seasonal nature [19] and certainly deadlier than the seasonal flu [20,21]. The higher-contagious Omicrons, which could override the immunity induced by both vaccines and prior infection [22] to cause breakthrough infection and reinfection, would continue to infect those who may be unvaccinated, vaccinated and boosted, even those with hybrid immunity, leading to hospitalization and death.

Although no longer a public health emergency, WHO still emphasized that COVID-19 remains a global health threat - the threat of “another variant” emerging to bring in new surges of disease and death, and of “another pathogen” emerging with even deadlier potential [23]. This threat implies two likelihoods: first, by natural evolution [24], a more pathogenic virus that takes a turn going beyond Omicron for a pandemic outbreak; and, second, by gain-of-function genetic manipulation [25,26] to create man-made recombinant variant able to infect systemically as a new bioweapon. Both scenarios could bring about an unthinkable COVID-catastrophe. The threat of bioterrorism could be very real as has been recently narrated [27].

Since immunity of current authorized vaccines would fade fast and so reinfection would not be uncommon. The question remains as to how the vaccine-elicited B cell and T cell immunity should be designed, generated and sustained. Aside from a plethora of known safety concerns, current authorized Spike-only vaccines would provide only limited, short-lived antibody protection against infection [28] without developing a fuller T cell immunity [29]. Instead of gaining immunity via infection, a vaccine platform able to induce safe, potent, broad and durable memory T cell immunity is highly desirable, to head off reinfection and long COVID altogether [30-35].

Of note, breakthrough infection or reinfection, despite the vast majority of the cases are asymptomatic and symptomatic mild illness as of this date [36,37], would add risks of mortality, hospitalization and other health hazards including burden of long COVID, in particular for the elderly, the immunocompromised, and who have co-morbidities [38,39].

It has been validated that viral Spike protein controls infectivity, while non-Spike proteins drive the virulency, i.e., disease severity and death [25]. Immune responses derived from vaccines targeting T cell epitopes on non-Spike proteins are pivotal in eliciting an upfront interferon response and memory immunity for viral clearance, which could lead to prevention of severe disease and ultimate reinfection [30-35]. Non-Spike proteins as immunogens have been literally overlooked in vaccine development since the outset of Warp Speed Operation three years ago [40]. For advance preparedness to avert and manage a potentially new catastrophic outbreak, an innovative genre of new next-generation vaccines has yet to be made in urgency.

We have demonstrated potent, broad, and durable T-cell and B-cell immunity of the pan-Sarbecovirus vaccine UB-612 in previous preclinical and clinical studies [29,41,42]. In the present observational clinical study, extended from a clinical Phase-2 trial conducted in Taiwan, we report a corresponding strong, long-lasting ≥10 month protection effect against COVID-19 moderate-severe disease (0% cases of hospitalization and ICU admission) by UB-612 universal vaccine, constructed with five designer peptides comprising sequence-conserved helper and cytotoxic T lymphocyte (Th/CTL) epitopes derived from Spike (S2×3), Membrane (M) and Nucleocapsid (N) proteins across Sarbecovirus species (**S1 Table**) and S1-RBD subunit protein (original ancestral strain).

## METHODS

### Design of Retrospective, Observational Study

#### Recruitment of study participants

This Observational Study (V-205-Q) was conducted in Taiwan as a retrospective, questionnaire-based survey that served an extension of the Phase 2 Clinical Trial of UB-612 vaccine in adolescent, younger and elderly adults (V205; ClinicalTials.gov ID: NCT04773067). A total of 3654 participants who had received UB-612 primary series were identified and invited to join the web-based questionnaire survey (**Fig 1**). The survey was conducted from October 2022 to March 2023. All eligible individuals were initially contacted by phone to ascertain study consent and provided with a hyperlink to access the web-based questionnaire.

**Figure 1.**
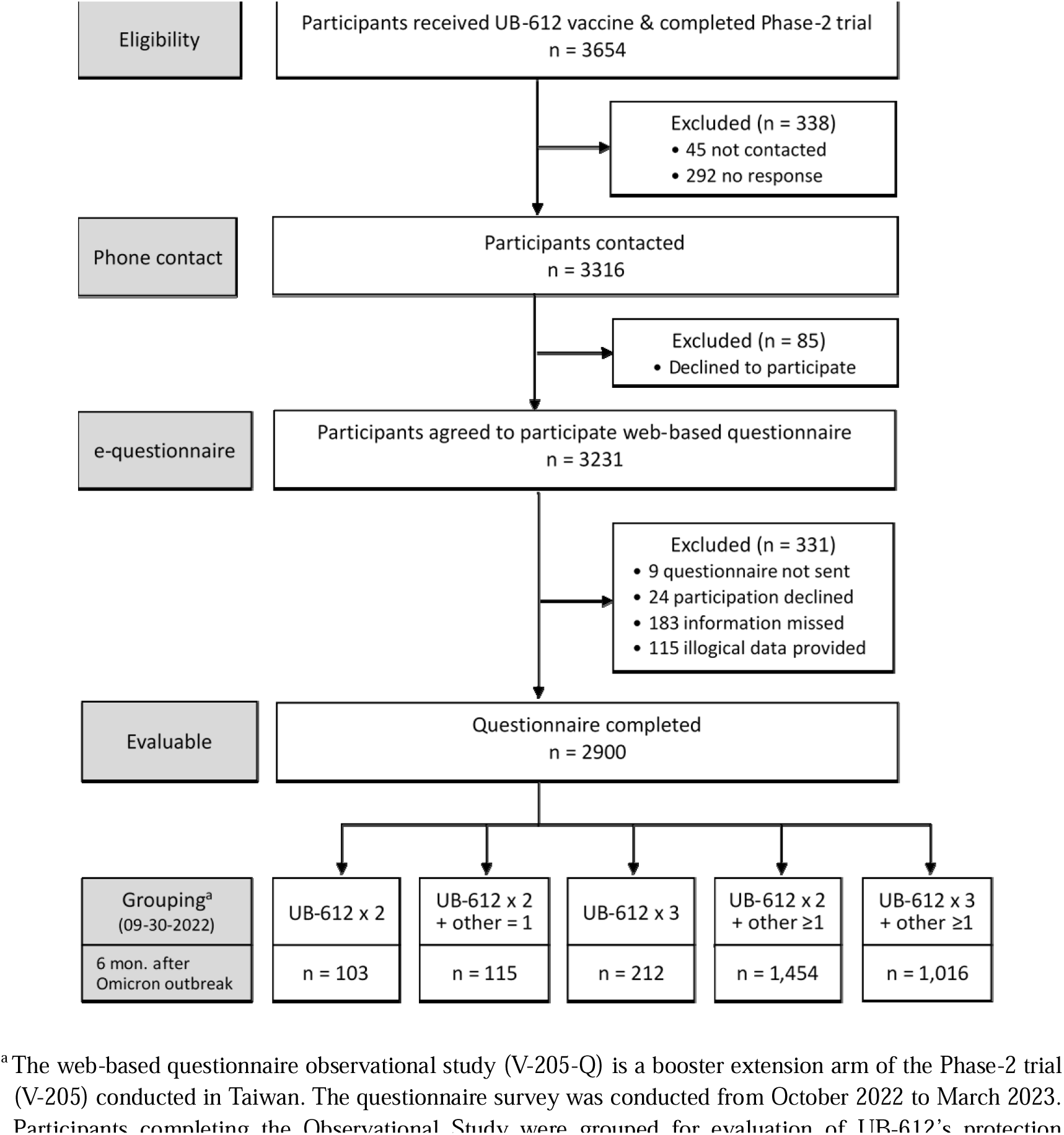
Flow chart of the questionnaire-based, retrospective Observational Study of UB-612 extended from the Phase-2 trial of primary and booster series.

#### Web-based questionnaire survey

An anonymous survey was included in a SMS message with hyperlink that directed participants to the Viedoc cloud-based platform. The questionnaire provided a disclosure of all the necessary information regarding the survey and a consent form on the first page. Once participants agreed to participate, they were able to continue with the rest of the questionnaire, which required them to provide information about their vaccination schedule, SASR-CoV-2 infection history and severity level. Additionally, any changes and repeatedly filling out of the questionnaire were prohibited after the completion of the survey to maintain the integrity of the data collected.

Two sources were used to obtain information on vaccination status and the history of SARS-CoV-2 infection. First, participants were requested to provide the dates of receiving COVID-19 vaccines regardless of types as recorded on their Vaccination Record Card. Second, the information on SARS-CoV-2 infection was obtained from the “COVID-19 Designated Residence Isolation (Home Isolation) Notice and Right to Petition for Habeas Corpus Relief,” which is given to individuals who test positive for COVID-19 through a rapid antigen test or reverse transcription polymerase chain reaction (RT-PCR) and are reported by the medical providers or self-report to the government.

#### Inclusion and exclusion of responses

Responses from study participants were reviewed for inclusion based on the completeness of the questionnaire. Invalid response was identified by the presence of one or more of the followings: (1) missing responses, (2) incorrect reporting of dates of vaccination dates and reports of SARS-CoV-2 infection within the phase 2 clinical trials of the UB-612 vaccine, and (3) reporting of vaccination dates that were not in chronological order.

#### Definitions of vaccination status and COVID-19 severity

The observation period was determined from the date of completing UB-612 primary series to 6 months after the Omicron outbreak in Taiwan, specifically until September 30, 2022, the date the study participants were categorized into five groups based on their vaccination status. These groups includes individuals who had received only UB-612 primary 2-dose series, received UB-612 primary series with a homologous booster, received UB-612 primary series with a heterologous booster, received the UB-612 primary series with more than one heterologous vaccines, and who received 3 doses of UB-612 vaccine with at least one heterologous vaccine.

Each participant was required to evaluate their situation based on the given definition to determine the severity level of COVID-19 disease. Asymptomatic cases were classified as individuals with COVID-19 who showed no symptoms. Those who experienced COVID-19 symptoms but did not require hospitalization fell into the mild case category. Moderate and severe cases were defined as individuals with COVID-19 requiring hospitalization and ICU admission, respectively.

### Vaccine immunity by humoral antibodies and cellular T cell responses

The UB-612’s booster protection effects against infection (asymptomatic and symptomatic mild cases) and moderate-severe disease may be revealed more in the post-booster neutralizing antibody strength and in cellular T cell immunity, respectively. We performed a random selection of 12 serum samples collected from participants who had received a third dose of the UB-612 vaccine to conduct live virus neutralization assay (VNT_50_) and pseudovirus neutralization assay (pVNT_50_) against SARS-CoV-2 wild type as well as relevant Omicron subvariants. Additionally, the relevant participants within the Observation Study with CD4 and CD8 T cell immunity previously profiled (ELISpot and ICS assays) were adapted for analysis of infection-vs.-T cell activity. The details of experimental procedures for assays of viral neutralization and T cell immunity are provided in the ***Supplemental Methods*.**

#### Statistics

Categorical variables were presented using counts and percentages, while continuous variables were presented as means with standard deviations, medians, and ranges. The Spearman’s rank correlation was performed to assess the relationship between the live virus neutralization assay (VNT_50_) vs. pseudovirus neutralization assay (pVNT_50_), VNT_50_ (WT) vs. T cell responses, and infection vs. T cell activity.

#### Study approval and ethics statement

The Observational Study (V-205-Q) adhered to the International Council for Harmonization of Technical Requirements for Pharmaceuticals for Human Use, Good Clinical Practice guidelines, Declaration of Helsinki and all relevant government regulations. The protocol, web-based questionnaire and informed consent form were approved by Taiwan Food and Drug Administration (TFDA) and the Institutional Review Boards (IRB) of the participating medical centers (see *Supplemental Appendices 1 and 2*).

It is important to note that this survey posed no more than minimal risk to the subjects involved. Eligible participants were selected from among those who volunteered to take part in the Phase 2 Clinical Trial. Investigators were approved to obtain verbal consent from participants over the phone by IRB during the screening process. Additionally, participants were requested to reaffirm their willingness after receiving the web-based questionnaire with a thorough explanation of the research. It was considered informed consent, if participants agree to participate after reading the information and fill in the questionnaire.

To ensure confidentiality throughout the survey, participants were de-identified by assigning to a coding number and were not required to provide identifiable information in the questionnaire. All data was collected via a cloud-based platform (Viedoc Technologies) and is accessible solely by the principal investigators, study coordinators at the sites, and authorized employees at StatPlus, Inc. (Contract Research Organization; CRO) and UBI Aisa (study sponsor).

## RESULTS

### Design of the Observational Study

Initial V-205-Q study enrollees (**Fig 1**) comprised young and elderly participants aged 12-85 years with approximately equal gender distribution (**Table 1**), who received “at least” UB-612 primary 2-dose series that included receiving one homologous booster [29,42] and heterologous booster(s). During the years 2020 and 2021 (pre-Omicron era), Taiwan successfully contained the spread of COVID-19 with very low infection rates reported, except only a blip of Alpha variant infection incidence confirmed in April-July 2021 (**Figure 2A)**.

**Figure 2.**
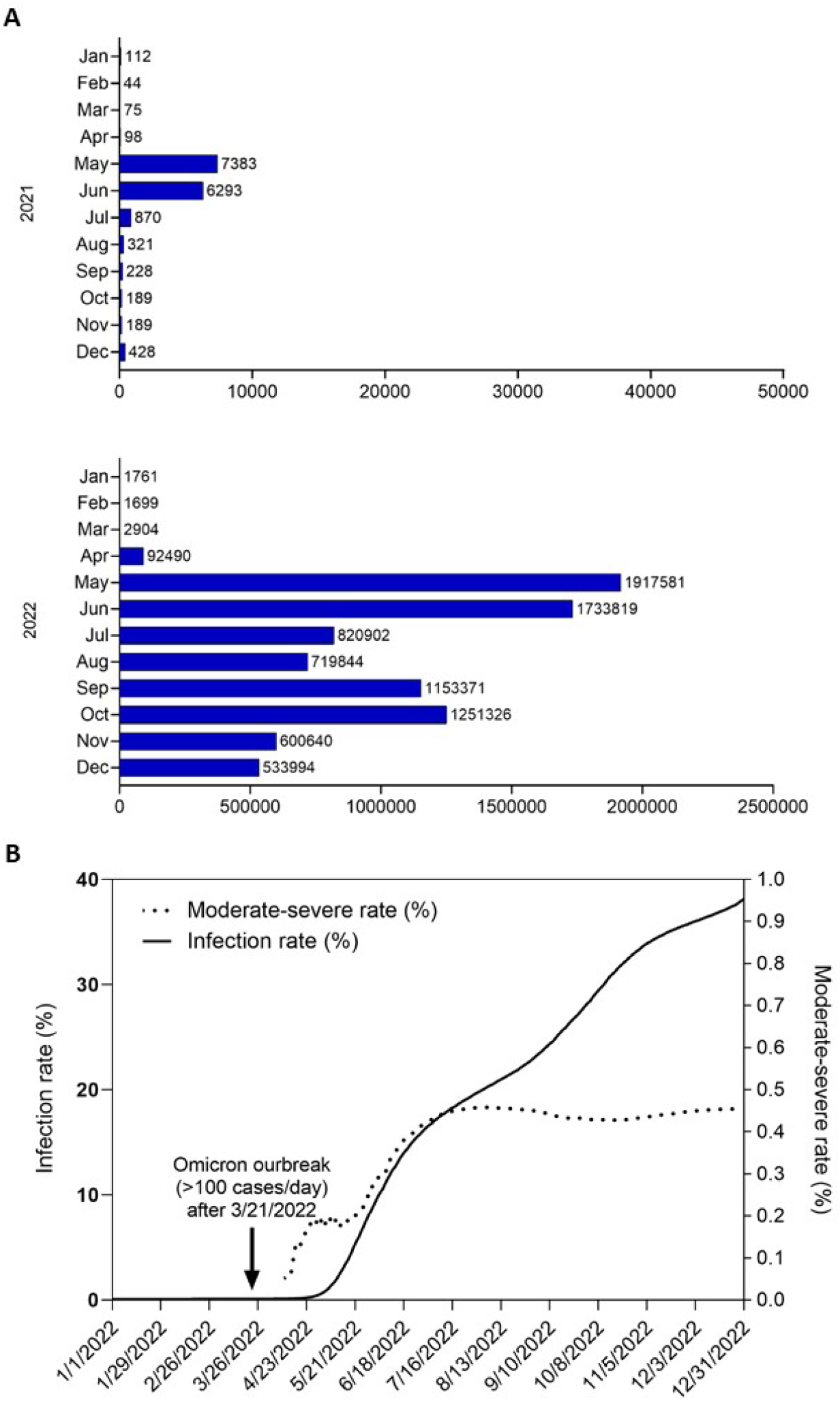
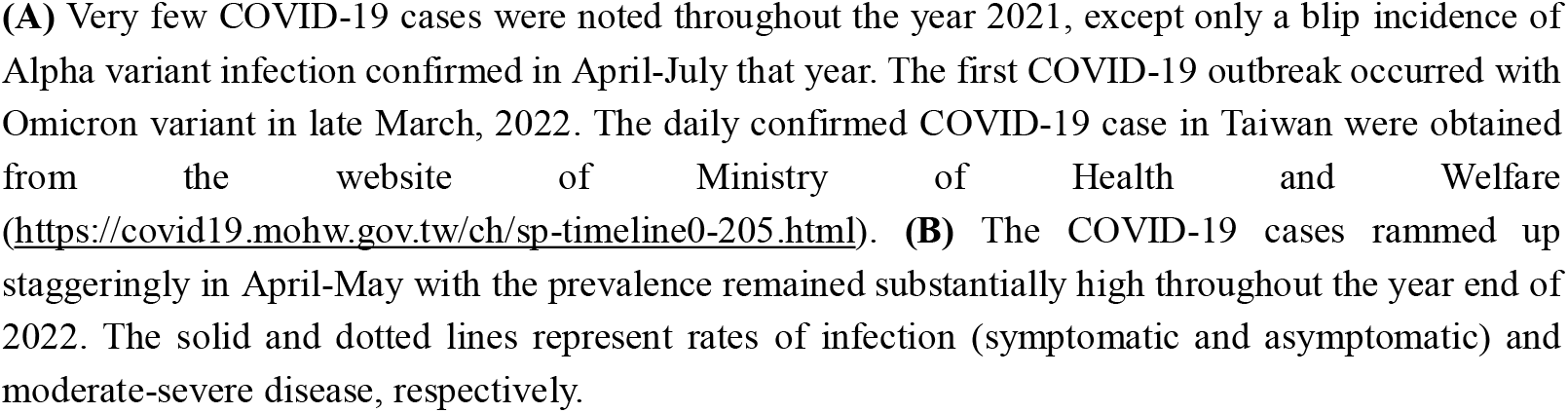
The 2022 outbreak of Omicrons and COVID-19 prevalence in Taiwan.

**Table 1.** Demograpgics of participants in the questionnaire-based Observational Study for evaluation of protection effect by UB-612 vaccine.

|  | UB-612 (homologous 2 or 3 doses, with or without heterologous booster) |  |  |  |  |  |
| --- | --- | --- | --- | --- | --- | --- |
|  | All subjects<br>(n = 2,900) | 2+0<sup>†</sup><br>(n = 103) | 2+1<sup>†</sup><br>(n = 115) | 3+0<sup>†</sup><br>(n = 212) | 2+X<sup>†</sup><br>(n = 1,454) | 3+X<sup>†</sup><br>(n = 1,016) |
| **Age<sup>‡</sup>** |  |  |  |  |  |  |
| Median | 40 | 31 | 17 | 44.5 | 38 | 43 |
| Range | 12-82 | 12-77 | 12-76 | 20-80 | 12-82 | 19-82 |
| **Age group<sup>‡</sup>** |  |  |  |  |  |  |
| 12-17 | 299 (10.3%) | 38 (36.9%) | 65 (56.5%) | 0 (0.0%) | 196 (13.5%) | 0 (0.0%) |
| 18-64 | 2,059 (71.0%) | 48 (46.6%) | 41 (35.7%) | 159 (75.0%) | 1,039 (71.5%) | 772 (76.0%) |
| ≥65 | 542 (18.7%) | 17 (16.5%) | 9 (7.8%) | 53 (25.0%) | 219 (15.1%) | 244 (24.0%) |
| **Sex** |  |  |  |  |  |  |
| Male | 1,476 (50.9%) | 68 (66.0%) | 63 (54.8%) | 114 (53.8%) | 691 (47.5%) | 540 (53.1%) |
| Female | 1,424 (49.1%) | 35 (34.0%) | 52 (45.2%) | 98 (46.2%) | 763 (52.5%) | 476 (46.9%) |
| **BMI (kg/m<sup>2</sup>)<sup>‡</sup>** |  |  |  |  |  |  |
| Median | 24.2 | 22.7 | 21.6 | 25.2 | 23.8 | 24.8 |
| Range | 13.1-50.9 | 15.9-44.7 | 15.6-35.9 | 15.8-42.9 | 13.1-50.9 | 16.7-50.1 |
| **BMI group, n (%)<sup>‡</sup>** |  |  |  |  |  |  |
| <30 kg/m<sup>2</sup> | 2,578 (88.9%) | 95 (92.2%) | 108 (93.9%) | 185 (87.3%) | 1,310 (90.2%) | 880 (86.6%) |
| ≥30 kg/m<sup>2</sup> | 321 (11.1%) | 8 (7.8%) | 7 (6.1%) | 27 (12.7%) | 143 (9.8%) | 136 (13.4%) |
| **Comorbidity, n (%)** |  |  |  |  |  |  |
| No | 2,838 (97.9%) | 101 (98.1%) | 115 (100.0%) | 208 (98.1%) | 1,423 (97.9%) | 991 (97.5%) |
| Yes | 62 (2.1%) | 2 (1.9%) | 0 (0.0%) | 4 (1.9%) | 31 (2.1%) | 25 (2.5%) |

<sup>†</sup> 2+0: individuals who had received only the UB-612 primary series; 2+1: individuals who had received the UB-612 primary series with a heterologous booster; 3+0: individuals who had received the UB-612 primary series with a homologous booster; 2+X: individuals who had received the UB-612 primary series with more than one heterologous vaccines; 3+X: individuals who had received 3 doses of the UB-612 vaccine with at least one heterologous vaccines.
<sup>‡</sup> Age and BMI were presented at the time of participating in the Phase-2 clinical trial of the UB-612 vaccine.

However, an outbreak occurred in late March 2022 due to a massive surge of highly-contagious Omicrons (**Fig 2A; S2 Table**). This surge period protracted with high infection incidence in local population which allowed a window of opportunity to assess clinical effect of those receiving UB-612 vaccine in our large Phase-2 and its extension studies for assessing breakthrough infection, mainly by Omicron BA.2 (BA.2.75 and XBB not present) and BA.5 (BF.7 and BQ.1 not present), observed through September 30, 2022, and analyzing the vaccine’s protection effects against COVID-19.

### Participants for evaluation of vaccine protection

The participants eligible for evaluation of vaccine effectiveness included those who received UB-612 primary 2-dose series only, UB-612 primary 2-dose series with 1 homologous booster, and UB-612 2-dose or 3-dose series plus 1 or more heterologous boosters during the observational phase up to September 30, 2022 (**Fig 1**). Of 2900 enrollees, the actual number of participants who stayed clean on the UB-612 course (primary 2-dose with or without 1 homologous booster) for protection evaluation was limited (**Table 1**), primarily due to a non-EUA approval status in Taiwan, for which the majority of vaccinees were referred to receiving heterologous booster over time (**Fig 1**), thus lowering the number of participants in completion of the scheduled course of primary 2-dose plus homologous 1-dose booster series.

### Protection against moderate-severe disease (hospitalization and ICU admission)

Six and 10 months post-booster vaccination, there were respective 337 (May 11, 2022) and 212 (September 30, 2022) participants who stayed clean on the homologous booster course (**S2 Fig**; **Table 3**). Protection effects of UB-612 vaccine against COVID-19 moderate-severe disease (hospitalization and ICU admission, H-ICU used hereafter) were evaluated during two critical time points, i.e., 6 months post-booster (May 11, 2022) and 6 months post-Omicron outbreak (September 30, 2022).

Of the 212 vaccinees on UB-612 booster, observed at ≥10 months after a homologous 3^rd^ dose, none has reported any case (0%) of H-ICU [43-45] (**Table 3**). Even 103 participants receiving only primary 2-dose series, all reported being free of H-ICU ≥14 months post-2^nd^ dose. No cases of H-ICU were reported in the 115 vaccinees who received primary 2-dose UB-612 plus 1 heterologous booster dose.

In contrast with 0% H-ICU rate per UB-612 vaccination, the overall Taiwan population on EUA-authorized vaccines reported inverse dose number-dependent H-ICU rates (moderate-severe disease), ranging from 0.44%, 0.22%, 0.11%, to 0.06% for individuals unvaccinated, receiving 1, 2, or ≥3 vaccine doses, respectively [43-45] (**Fig 3A**). These suggest that the unvaccinated people were more prone to contract severe COVID-19 following Omicron infection, and that booster vaccination may provide additional protection against moderate-severe disease.

**Figure 3.**
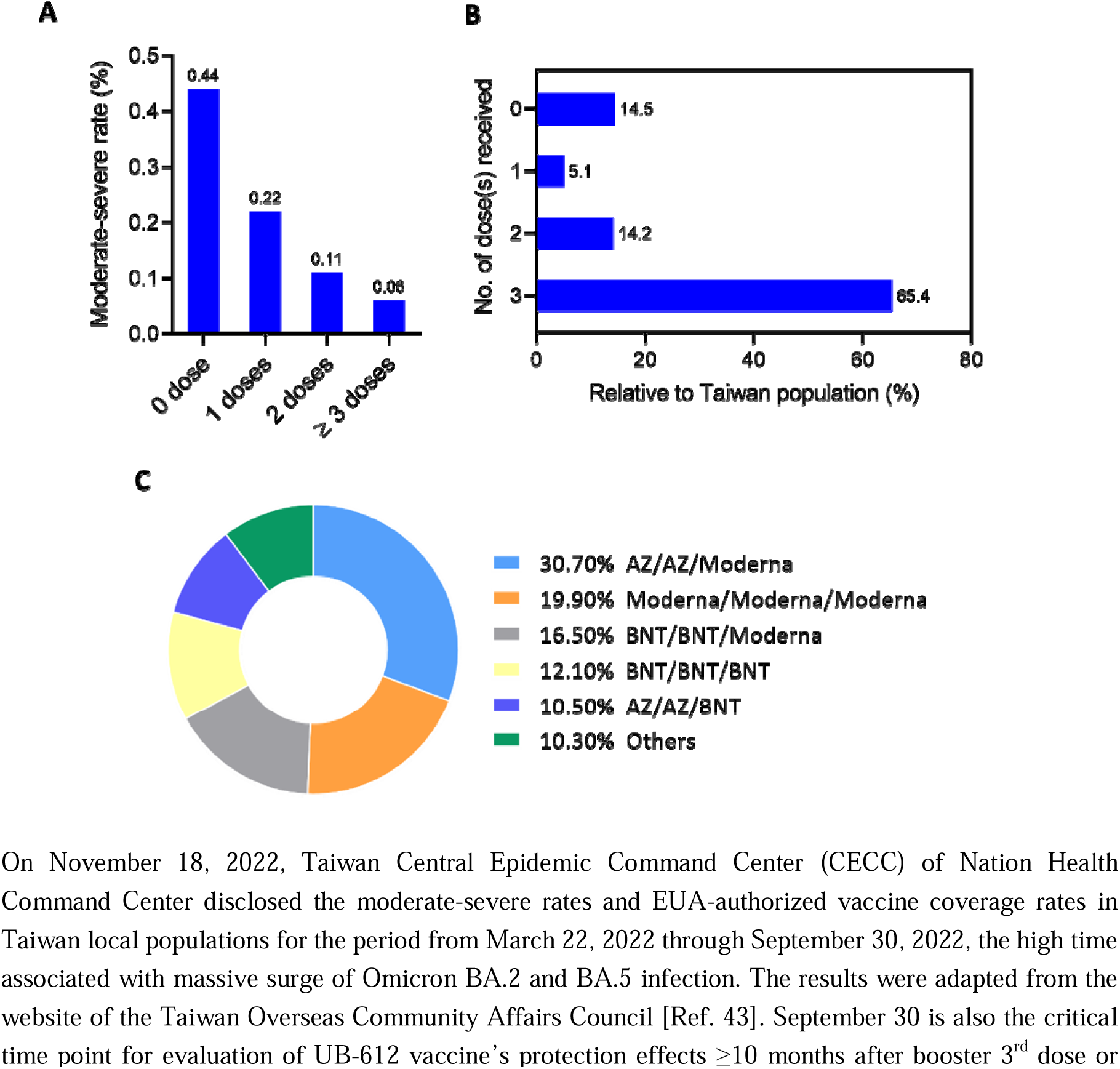
COVID-19 moderate-severe rate and vaccine coverage in Taiwan reported September 30, 2022.

At September 30, 2022, an overall infection rate of ∼30% and moderate-severe disease rate of 0.46% were recorded (**Fig 2B**) in Taiwan population while 65.4% of the population have received 3 doses or more of EUA-vaccines (**Fig 3B**), mainly AZD1222, BNT162b2 and mRNA-1273 (**Fig 3C**). Over the time course since Omicron outbreak from March 2022 to December 2022, the infection rate continued to increase, while the moderate-severe disease rate initially rammed up in parallel with infection rate (**Fig 2B**), but shortly reached the peak at 0.47% in July and momentarily plateaued out through the year end.

### Protection against infection (asymptomatic and symptomatic mild disease)

Upon UB-612 booster vaccination during the BA.5 dominance period, we observed only a 1.2% infection rate (symptomatic and asymptomatic) at May 11, 2022 (6 months post-booster; i.e., 6 weeks after Omicron outbreak) (**Table 4A**), which increased to 27.8% at September 30, 2022 (observed ≥10 months post-booster; i.e. 6 months after Omicron outbreak).

By contrast, use of EUA-vaccines was associated with a higher 2.2% infection rate (symptomatic and asymptomatic) observed at May 11, 2022 (6 weeks after Omicron outbreak) in overall Taiwan population [44,45] (**Table 4B**), which increased up to an approximately same rate of 27.9% observed at September 30 (6 months after Omicron outbreak). At both time points, details about the vaccination status and the time elapse associated with EUA-vaccine populations in Taiwan were not disclosed by Taiwan CDC.

The infection rate in Taiwan population (unvaccinated and vaccinated) continued to climb up to ∼40% at the end of the year 2022 (**Fig 2B**), which was underestimated as the number did not cover those who were infected but unreported, including symptomatic and asymptomatic. Of note, during the same time course, people contracted COVID-19 moderate-severe disease started from 0% rate in late March that peaked at 0.46% overall in mid-July, and plateaued throughout the year end (**Fig 2B**), which is close to 0.44% observed for the unvaccinated (**Fig 3A**).

### Vaccine immunity behind protection effect

Along with the UB-612 Observational Study, we re-analysed the booster-recalled serum viral-neutralizing antibody titers that covered the increasingly higher-contagious Omicrons BA.5 and beyond (BA.5/XBB.1.5/BQ.1/CH.1.1) under limited source of serum samples. The booster memory T cell responses profiled previously were adapted to explore the better lead that might account for the observed vaccine protection effects.

#### Humoral viral-neutralizing antibodies

*Against live virus.* Consistent with the trend of live virus-neutralization assay results (VNT_50_ titers against Omicron BA.1/BA.2 at 670/485) observed from Phase-1 booster vaccination study participants (aged 20-55 yrs.) (29), UB-612 in the Phase-2 booster study participants (18-85 yrs.) induced similar, substantial amount of VNT_50_ titers of 359/325/123 against heavily mutated, immune-evasive Omicrons BA.1/BA.2/BA.5, respectively (**Fig 4A**). The values appear to far exceed those reported with other vaccine platforms, e.g., for anti-BA.1 VNT_50_ at a range of 46.2-106 (**S3 Table**); the corresponding VNT_50_ values by EUA vaccines against BA.2 and BA.5 have not been available, most likely due to very low levels beyond quantification.

**Figure 4.**
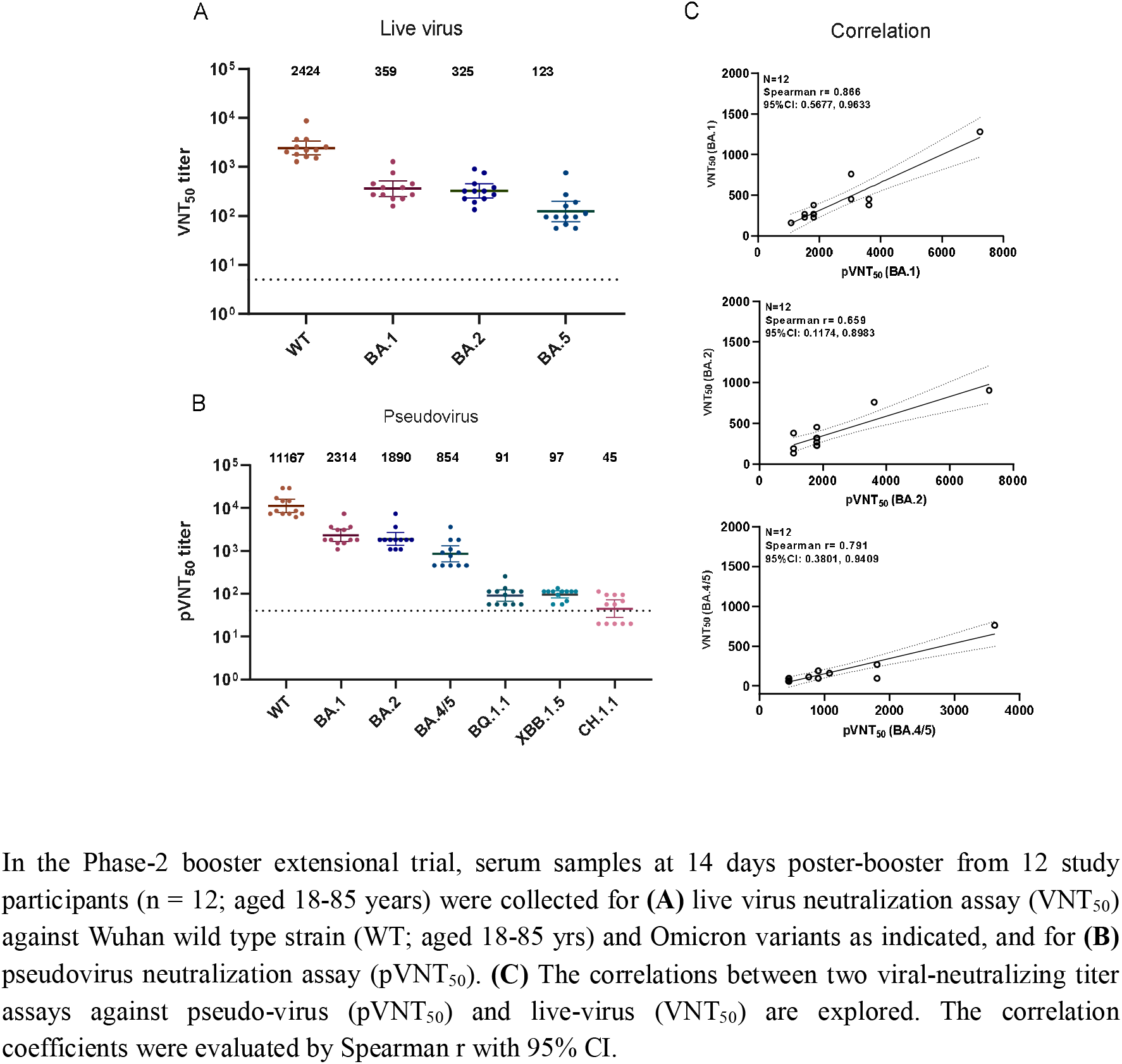
Viral-neutralization antibody titers and functional correlations between live virus VNT_50_ and pseudovirus pVNT_50_.

*Against pseudovirus.* We have shown previously that the Phase-2 UB-612 booster vaccination (3 homologous doses) bears a competitive edge over other vaccine platforms in regard to pseudovirus-neutralizing activities (pVNT_50_) against original wild type WT/BA.1/BA.2/BA.5 strain [29]. In a recent fresh viral-neutralization run against new subvariants, we found that there is a sharp decrease in pVNT_50_ by multiple folds for the variants beyond BA.5. The induced pVNT_50_ titers against BA.2/BA.5/XBB.1.5/BQ.1.1/CH.1.1 were at 1890/854/97/91/45, respectively (**Fig 4B**). The titer values appear to be higher than the previously reported counterpart titers of 475/197/59/25/8 from the 3-dose monovalent mRNA vaccine, and comparable to the counterpart titers of 2,151/1,099/241/86/66 from 3-dose monovalent + 1-dose bivalent mRNA vaccine [46].

A cliff drop in pVNT_50_ from BA.5 to XBB.1.5 is evident in another study that reported a 10-fold lower pVNT_50_ for XBB.1.5 as compared to BA.5 [47]. XBB.1.16 is 1.2-fold higher in immune-evasion. As stated above, the CH.1.1 variant is estimated to be 2- to 7-fold lower than XBB.1.5 in terms of pVNT_50_ level. These observations suggest that the neutralizing antibody titer strength is an increasingly a less relevant immunity parameter.

Per UB-612 vaccination, the live virus and pseudovirus neutralization assay results are functionally well correlated (**Fig 4C**), as exemplified by the assays against Omicron BA.1, BA.2, and BA.5 with a Spearman r = 0.866, 0.659, and 0.791, respectively. Of note, live virus-neutralizing titer VNT_50_ would make much better sense than pseudovirus-neutralizing titer pVNT_50_, as the latter assay is based on the interaction with viral outer Spike protein only. For other vaccine platforms, live virus-neutralizing VNT_50_ titer values beyond against Omicron BA.1 have not been reported (**S3 Table**).

### Cellular Interferon and cytotoxic T cell responses

UB-612 booster vaccination has been demonstrated previously to induce potent and durable Th1-oriented IFN-γ^+^-ELISpot responses (SFU/10^6^ PBMC) of 374/261/444 at the peak post-2^nd^ dose/pre-boost/peak post-booster, respectively (**S3A Fig**), along with a robust presence of cytotoxic CD8^+^ T cells frequency (peak post-2^nd^ dose/pre-boost/peak post-boost CD107a^+^-GranzymeB^+^ CD8 T cells, at 3.6%/1.8%/1.8%) (**S3B Fig**) (29). UB-612 appears to trigger (at stimulation with Th/CTL+RBD) far greater IFN-γ^+^-T cell ELISpot responses than those produced by the current Spike-only vaccines (**S3 Fig**).

In further analysis with T cell response data from available participants (4 infected and 3 not infected) involved in the Observational Study who received 3-dose UB-612 and reported moderate-severe disease free, the moderate-severe infection was found to be in high correlation with IFN-γ^+^ and CD107a^+^-GranzymeB^+^ cytotoxic CD8 T cells as biomarkers (**Fig 5A and 5B**), but not with IFN-γ^+^-CD4 (**Fig 5C**), as shown by the Spearman r = -0.728, -0.721, and -0.218, respectively. This indicates the multitope-memory killer CD8 T cells upon stimulation (infection or booster vaccination) by virus-specific antigens may play a pivotal role in protection against severe disease and reinfection.

**Figure 5.**
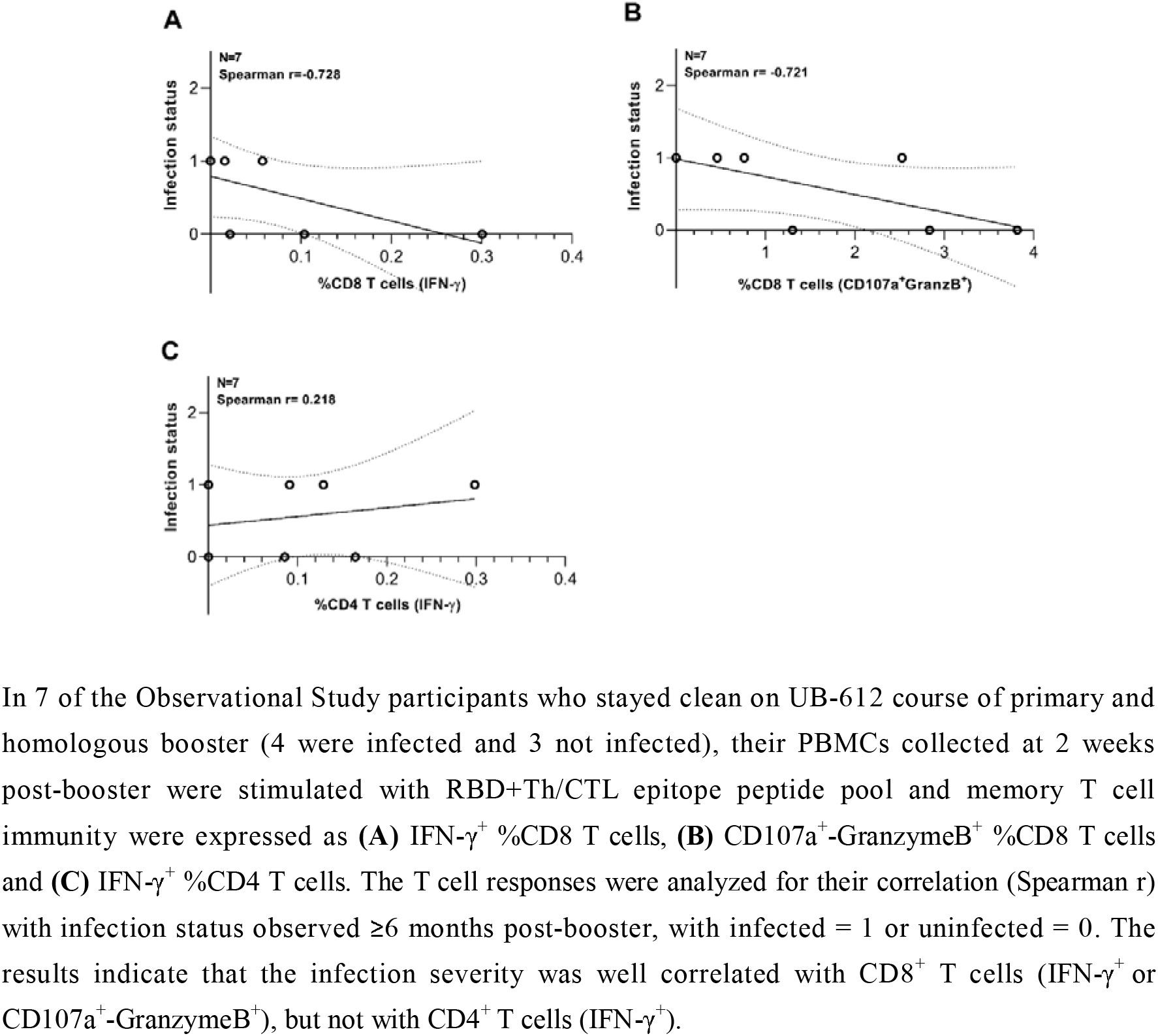
Cytotoxic CD8^+^ T cell responses induced by UB-612 booster vaccination correlate well with infection status.

## DISCUSSION

Conventionally, vaccine protection effectiveness or efficacy has been analyzed at 1-6 months post-vaccination [48-50]. However, the present UB-612 Observational Study faced hurdles of following such assessment paradigm for evaluation of protection effect, as Taiwan reported very low COVID 19 incidence prior to April 2022 (**Fig 2A; S2 Table**). Study participants received the 2^nd^ vaccine dose between March 2021 – August 2021, and the booster 3^rd^ dose between October 2021 and December 2021. The Omicron outbreak peaked around May 11, 2022, when 6 months of time had passed after UB-612 booster was administered (**Table 2**).

**Table 2.** Critical time points of the Observational Study V-205-Q in UB-612 Phase-2 trial V-205.

| Clinical Trial/Study | Date | Events<sup>a</sup> |
| --- | --- | --- |
| V-205 | 03/23/2021 - 08/21/2021 | **Received the 2<sup>nd</sup> dose (primary series)**<br>28 days post-1<sup>st</sup> dose |
| V-205 | 10/16/2021 - 12/09/2021 | **Received the 3<sup>rd</sup> dose (booster)**<br>~6.0 to 8.0 months post-2<sup>nd</sup> dose |
| V-205<br>V-205-Q | 12/23/2021 | **Two weeks after the 3<sup>rd</sup> dose**<br>V-205-Q observation initiated |
| V-205 | 02/08/2022 - 03/08/2022 | V-205 Follow-up completed |
| N/A | 03/21/2022 | **Omicron outbreak (≥100 cases/day)** |
| V-205-Q | 05/11/2022 | **Observation time-point 1 of V-205-Q**<br>Median day of 6 months after 3<sup>rd</sup> dose<br>Six weeks after Omicron outbreak |
| V-205-Q | 09/30/2022 | **Observation time-point 2 of V-205-Q**<br>Six months after Omicron outbreak<br>Ten months after the 3<sup>rd</sup> dose; 14 months after the 2<sup>nd</sup> dose |
| V-205-Q | 10/13/2022 | **The end of observation period**<br>The first questionnaire completed |

<sup>a</sup> V-205-Q is an IRB-approved, questionnaire-based, retrospective extension study from the Phase-2 trial (V-205) involving nearly 4,000 healthy participants who received primary 2 doses with 28 days apart and a booster (3<sup>rd</sup> dose) 6-8 months post-2<sup>nd</sup> dose [Refs. 29,42]. All participants of this extension study (V-205-Q) had to receive at last the primary two doses of UB-612. The time periods participants received the 2<sup>nd</sup> and the 3<sup>rd</sup> dose of UB-612 were for March-August 2021 and October-December 2021, respectively. The one year follow-up of V-205 trial was completed from February 8 to March 8, 2022. The start and end of observing period in this study was defined as the last subjects received 3<sup>rd</sup> doses of UB-612 after two weeks and the first questionnaire completed, respectively. The Omicron outbreak in Taiwan is defined by the median day of 7-days average >100

As the Phase-2 booster dose was administered 6 to 8 months after the 2-dose primary series [29], we elected to evaluate the booster effect at 6 months after Omicron outbreak (September 30, 2022) that laid path to revelation of a rough annual prevention outcome. This marks a time elapse of ≥10-12 months after UB-612 booster vaccination, corresponding to ≥14-18 months post-2^nd^ dose for participants who received only the UB-612 primary 2-dose series without taking homologous or heterologous booster.

The protection effects by UB-612 vaccination are strong and long-lasting. Overall, none (0%) of COVID-19 H-ICU cases (moderate-severe disease) were observed over 12 months (**Table 3**) – which would mean much more clinical significance than the protection outcomes garnered conventionally within 90 days or up to 6 months post-vaccination – despite a small yet significant sample size for observational analysis (n = 212 for those on 1 homologous booster dose and n = 103 for those on 2-dose primary series without any further dose).

**Table 3.** COVID-19 disease severity observed ≥10 months post-booster (3^rd^-dose) and ≥14 months post-2^nd^ dose of UB-612 vaccine in the Phase-2 extension Observational Study.

| Characteristics |  |  | COVID-19 Severity<sup>a</sup> |  |  |
| --- | --- | --- | --- | --- | --- |
|  |  |  | Asymptomatic (%)<sup>b</sup> | Mild (%)<sup>b</sup> | Moderate-Severe (%)<sup>b</sup> |
| Vaccination status |  |  |  |  |  |
| UB-612 | non-UB-612 | Total (%) |  |  |  |
| 2 | 0 | 103 (3.6%) | 0 (0.0%) | 32 (31.1%) | 0 (0.00%) |
| 3 | 0 | 212 (7.3%) | 4 (1.9%) | 55 (25.9%) | 0 (0.00%) |
| 2 | 1 | 115 (4.0%) | 1 (2.8%) | 35 (30.4%) | 0 (0.00%) |
| 2 | $\geq 1$ | 1,569 (54.1%) | 18 (1.1%) | 453 (28.9%) | 2 (0.13%) |
| 3 | $\geq 1$ | 1,016 (35.0%) | 16 (1.5%) | 254 (25.1%) | 2 (0.20%) |

<sup>a</sup> Per UB-612 vaccination, the outcomes of infection rate and disease severity, observed on September 30, 2022 (i.e. $\sim \geq 10$ post-booster; $\sim \geq 14$ months post-2<sup>nd</sup> dose).
<sup>b</sup> Percentage = (Value/Total) × 100%.

Of further interest, no cases of H-ICU (0% rate) were recorded, either, in the 115 vaccinees who received primary 2-dose UB-612 plus 1 heterologous booster dose (**Table 3**), but not for those receiving UB-612 (2 and 3 doses) plus more than 1 heterologous booster, who reported H-ICH rates of 0.13% and 0.20%, respectively, for reasons unknown.

The overall Taiwan populations of the unvaccinated and those received EUA-authorized Spike-only vaccines of any vaccination status were recorded with a neat inverse dose number-dependent rate of H-ICU that ranged from 0.44, 0.22, 0.11 to 0.06% (**Fig 3A**). The clear additional protection benefit against disease severity during Omicron surge span is consistent with the previous report on booster vaccination [2].

In addition to S1-RBD-focused targeting that elicits decent levels of viral-neutralizing antibodies up to BA.5 variant (**Fig 4**), the distinct difference in protection against H-ICU between UB-612 and Spike-only vaccine platforms could be accounted for by the fact that UB-612 also targets multiple conserved Th/CTL epitopes on both Spike and non-Spike proteins (**S1 Table**), leading to a robust, durable memory T cell immunity that is universally multiple recognizing (**S3 Fig**). This could be essential to limiting the impact of rapid viral mutation, and is likely to serve as a pillar for high protection against severe disease by the increasingly immune-evasive Omicron variants [33-35].

UB-612 induces roust, durable T cell responses (**S3 Fig**) that may combat Sarbecovirus variants equally as the vaccine targets conserved (universal) Th/CTL epitopes on Spike and non-Spike proteins. As an infection-induced immunity by all SARS variants would involve targeting Spike and non-Spike proteins, the UB-612-induced immunity would be substantially closer (vs. Spike-only vaccine platforms), if not mimicking, to that induced by natural infection. A hybrid immunity associated with UB-612 vaccination as either primer or booster may provide potent protection effects against severe reinfection.

That 0% of H-ICU cases observed ≥10 months after UB-612 primary or booster series (**Table 3**) suggests that the vaccine-induced immunity and the associated protection against H-ICU may be strong and long-lasting. This appears to be similar to that by infection immunity in a recent large meta-analysis study showing that the vaccine-naïve natural immunity of post-Omicron BA.1 infection exhibits prolonged, high protection (88.9%) against severe disease for at least 40 weeks [51], which wanes very slowly. With respect to symptomatic mild disease, the vaccine-naïve BA.1-infection immunity at weeks 40, by estimation, provides ∼40% effectiveness [51]. UB-612 vaccination exhibits an infection rate of 27.8% observed at weeks 40 (September 30, 2022; ≥10 months post-booster) (**Table 4A**). It should be noted that the UB-612’s effects against moderate-severe disease and symptomatic infection are not equivalent to protection effectiveness by definition.

**Table 4.** Infection rates of COVID-19 observed on two specific dates in populations receiving EUA-vaccines in Taiwan.

**A. Infection rate in study participants receiving UB-612 vaccine doses<sup>a</sup>**
| Observation time point | Date | Total samples | Cases | Infection rate |
| --- | --- | --- | --- | --- |
| 6 mon. after 3<sup>rd</sup> dose<br>(6 wks after Omicron outbreak) | 05/11/2022 | 337 | 4 | 1.2% |
| 10 mon. after 3<sup>rd</sup> dose<br>(6 mon. after Omicron outbreak) | 09/30/2022 | 212 | 59 | 27.8% |

**B. Infection rate in Taiwan population on EUA-authorized vaccine doses<sup>b</sup>**
| Observation time-point | Date | Taiwan population | Cases | Infection rate |
| --- | --- | --- | --- | --- |
| 6 wks after Omicron outbreak | 05/11/2022 | 23,198,133 | 505,455 | 2.2% |
| 6 mon. after Omicron outbreak | 09/30/2022 |  | 6,461,400 | 27.9% |

<sup>a</sup> For UB-612 booster, infection rates on the median date after 6 months post-booster (05/11/2022) and 10 months post-booster (09/30/2022, the date 6 months after Omicron outbreak).
<sup>b</sup> Infection rates observed on the same two dates for the populations in Taiwan receiving EUA-authorized vaccines. Data from: Taiwan Central Epidemic Command Center (CECC) of Nation Health Command Center and the websites of National Center for High-performance Computing (NCHC), Taiwan [Ref. 44]; the website of Department of Household Registration, Taiwan [Ref. 45].

Moreover, the notable protective effects against moderate-severe disease and symptomatic infection through UB-612 vaccination (2 or 3 doses) bears a similarity as well to that reported by hybrid immunity. The effectiveness of hybrid immunity (vs. infection immunity) against hospital admission or severe disease was 97.4% (vs. 74.6%), and that against reinfection was 41.8% (vs. 24.7%) at 12 months after primary vaccination [52]. Similar hybrid immunity’s protection effectiveness rates (against reinfection and severe disease) were noted after vaccine booster third dose in the same study. Hybrid immunity provides clearly better protections than by plain natural infection immunity.

In the aforementioned meta-analysis on protection against post-BA.1 symptomatic and severe diseases [51], relative to those by the vaccine-naïve natural immunity, the mRNA and adeno-vectored vaccines were shown to be substantially less effective and fading faster. This is consistent with an earlier study [53] revealing that prior natural infection was associated with lower incidence of symptomatic and severe reinfection, compared to mRNA vaccines.

Aside from being less effective, short-lived protection efficacy against infection [28], the “monovalent” mRNA vaccine’s protection effectiveness against severe disease and death through booster vaccination is being challenged [54]. On the other hand, the “bivalent” mRNA vaccine as booster has been shown in all age groups to provide a peak ∼60% effectiveness at day 15 against severe Omicron infection and wane over 99 days [50]; and, in the elderly (>65 years), only a 72% effectiveness against hospitalization was observed at a short period of 120 days [55], with a longer-term outcome remained to be investigated. Of concerns as reported during the BA.4/5-, BQ-, and XBB-dominant phases, the higher the number of monovalent mRNA vaccine previously received, the higher the risk of contracting COVID-19 upon bivalent vaccination [56].

While natural immunity and hybrid immunity would provide high, long-lasting severe disease-protecting effectiveness, a reinfection of any degree would add on risks of mortality, hospitalization and other health hazards including burden of long COVID [38,39]. Of high concerns, there has been an increasingly high incidence of Omicron-to-Omicron reinfections over a shorter time interval than that seen in pre-Omicron Variants of Concern [57].

Several limitations should be acknowledged in relation to the e-questionnaire survey employed in this study. First, the possibility of bias arising from recall error or misreporting cannot be entirely ruled out, though we did exclude participants who made errors in chronological order of the immunization schedule. Second, due to a non-EUA approval status, volunteers were directed to receive only EUA-approved vaccines during the Omicron outbreak, resulting in challenges in collecting a large, representative sample size, particularly for the groups who had completed the UB-612 primary series only or received a homologous booster. Third, as a questionnaire survey, COVID-19 cases were limited to those participants who had received a “COVID-19 Home Isolation Notice”, which could only be obtained if they had either tested positive and been reported by healthcare providers or had self-reported to local authorities. Hence, the cases observed in this study might have been underestimated.

UB-612 vaccine has been demonstrated to be safe and well tolerated without concerns of severe adverse events identified in our Phase-1 and Phase-2 clinical trials [29,42]; and to significantly reduce viral loads, lung pathology scores, and disease progression in mice and macaques [41]. The design with conserved Th/CTL epitope peptides produces broadly recognizing, potent and durable T cell responses (**S3 Fig**), which may energize B cell humoral immunity with substantial “live” virus-neutralizing antibodies up to BA.5 subvariant (**Fig 4A**). Further, preliminary results revealed in a Phase-3 study [ClinicalTrials.gov NCT05293665] [58] suggest UB-612 may be a competent heterologous booster to mRNA, adenovector-based and virus-inactivated vaccine platforms as UB-612 can enhance their viral-neutralizing titer and seroconversion rate against Omicron BA.5 subvariant (**S3 Table**).

Beyond BA.5, we have seen from UB-612 booster sera a generally-known pattern of cliff drop in pseudovirus-neutralizing titer against the extraordinarily immune-evasive BQ.1.1/XBB.1.5, and CH.1.1 subvariants [46,47], close to 10- and 20-fold reduction relative to BA.5, respectively, as shown in the present study (**Fig 4B**). In face of antibody-evading mutants and a growing number of people getting reinfected [57,59], the observations in the present report suggest that viral-neutralizing antibody titer strength may be increasingly becoming a less relevant immunity parameter than the memory T cell immunity in regard of providing long-term control of reinfection, hospitalization, and severe disease. Further, cytotoxic CD8 T cell immunity induced by UB-612 vaccination (**Fig 5**) is suggestive that it may be the ultimate pivot that controls the infection disease severity. Of note, potent memory CD4 and CD8 T cell immunity may protect against SARS-CoV-2 infection in the absence of immune neutralizing antibodies [60,61].

Instead of resorting to frequent booster jab or variant-updated vaccines, one of the pragmatic approaches to curbing immune-evasive impacts on COVID-19 disease severity would be the exploration of universal, pan-coronavirus like vaccines, which has been strongly advocated [62,63]. UB-612 represents a “pan-Sarbecovirus vaccine” as it is constructed with multiple sequence-conserved, non-mutable Th/CTL epitopes on Spike and non-Spike proteins across all viral variants (**S1 Table**). While non-Spike proteins (e.g., E, M and N) are critical for host interferon response and memory T cell response [30-35], they can mutate as well (**S4 Table**). In addition to antibody-evasion, mutations on Spike and non-Spike could lead to T cell escape [64,65].

The pack of non-Spike proteins, but not Spike protein, has been neatly confirmed to pivot the COVID-19 disease severity and mortality [25]; and, so, a resurgence of deadly variants can be readily made possible through the gain-of-function genetic manipulation [24,25] to infect systemically (as a new bioweapon) causing new COVID-19 outbreak. The concerns over the mutation on the pathogenicity-determinant non-Spike proteins and the likelihood of new pandemic catastrophe underscore the essentiality and urgency of incorporating conserved T cell epitopes in the making of better next-generation COVID-19 vaccines.

The observation of zero cases of moderate-severe COVID-19 disease (**Table 3**) may be attributable to UB-612’s multiple immunogens targeting Spike (S1-RBD and conserved sites on S2) and non-Spike targets (conserved sites on M and N), in particular those sequence-conserved and non-mutable Th/CTL lymphocyte epitopes. While the roles of T cell immunity in protection against severe disease and reinfection has been underestimated [30-35], there has been an optimistic trend on the horizon moving towards development of a universal mRNA T-cell vaccine (combination with BA.1- or BA.5-containing bivalent vaccine) that targets conserved epitopes on non-Spike proteins [66,67], aiming to enhance breadth of T cell immunity and lengthen duration of antibodies against severe disease and hospitalization.

Conceivably, the unique design of pan-Sarbecovirus, T cell immunity-promoting UB-612 vaccine (**Supplemental Figure 3**) by itself may lend a support to the finding of strong (0% of H-ICU cases) and long-lasting (≥10 months) protection against COVID-19 moderate-severe disease (**Table 3**). UB-612 vaccine, distinctly different from the contemporary Spike-only vaccine platforms, may play a significant role as a competent primer and booster. The ultimate protection effects and scientific insights against severe disease, amid the wild run of ever-emergent higher-contagious SARS-CoV-2 mutants, warrant further clinical investigations, including the impact on long COVID.

## Data Availability

All data produced in the present work are contained in the manuscript

## AUTHOR CONTRIBUTIONS

CYW and WJ were responsible for vaccine development including study protocol design and implementation of the clinical studies. CYW, WJ and BSK were responsible for interpretation of the clinical data. YHL, YHH YHP, HCC and LFF were responsible for data acquisition, analysis and preparation of respective report. YTY was responsible for management of laboratory testing and data preparation. CYW, YHL, YHP and BSK had full access to and verified all the data in the study and take responsibility for the integrity and accuracy of the data analysis. BSK and CYW drafted, prepared and reviewed the manuscript. All authors reviewed and approved the final version of the manuscript. CYW had final responsibility for the decision to submit for publication.

## DATA SHARING

The Observational Study protocol V-205-Q is provided in the *Supplemental Appendices*. All relevant data are within the manuscript and its *Supplemental Information*.

## DECLARATION OF INTERESTS

CYW is co-founder and board member of UBI, United BioPharma, and UBI Asia, and named as an inventor on several patent applications filed covering COVID vaccine development. WJP is also named as co-inventors on related patent applications. CYW, WJP, BSK, YHL, YHH, YHP, YTY, HCC and LFF are employees within the UBI group.

## ACKNOWLEDGEMENTS

The study was funded by UBI Asia (study sponsor) and the Taiwan Centers for Disease Control, Ministry of Health and Welfare. Team members at Institute of Biomedical Sciences, Academia Sinica for the live virus neutralization assay; and team members at the RNAi Core Facility, Academia Sinica for the live virus pseudovirus neutralization assay. All health convalescent sera were supplied by Biobank at the National Health Research Institutes (NHRI), Taiwan.

We thank all the trial participants for their dedication to these trials; the clinical investigators and investigation staff at China Medical University Hospital, Taipei Medical University Hospital, Far Eastern Memorial Hospital, National Cheng Kung University Hospital, Linkou Chang Gung Memorial Hospital, Kaohsiung Chang Gung Memorial Hospital, Kaohsiung Medical University Hospital, Tri-Service General Hospital, Taipei Veterans General Hospital, Kaohsiung Veterans General Hospital, Changhua Christian Hospital and Taichung Veterans General Hospital for their involvement in conducting the trial; and members of the IDMC for their dedication and guidance.

Special administrative support by Fran Volz from the UBI group is also acknowledged with gratitude.

## Notes

### Competing Interest Statement

The authors have declared no competing interest.

### Author Declarations

All IRB Approvals are located in the Supplemental Appendix 2. IRB Approval Tracker: Ethical Committee Endorsements for the Questionnaire-based, Retrospective, Phase-2 Extension Study

