## Supplemental Information for "Targeting Multiple Conserved T-Cell Epitopes for Protection against COVID-19 Moderate-Severe Disease by a Pan-Sarbecovirus Vaccine"

**Table of Contents** **Page**

#### Supplemental Methods

**Live virus-neutralization assay against wild type, BA.1, BA.2, and BA.5 variants.**

Neutralizing antibody titers were measured by CPE-based live virus neutralization assay using Vero-E6 cells challenged with wild type (SARS-CoV-2-Taiwan-CDC#4, Wuhan) and Omicron variant (SARS-CoV-2-Taiwan-CDC#16804, BA.1; SARS-CoV-2-Taiwan-CDC#19380, BA.2; SARS-CoV-2-Taiwan-CDC#689423, BA.5), which was conducted in a BSL-3 lab at Academia Sinica, Taiwan. Vero-E6 (ATCC CRL-1586) cells were cultured in DMEM (Hyclone) supplemented with 10% fetal bovine serum (FBS, Gibco) and 1x Penicillin-Streptomycin solution (Thermo) in a humidified atmosphere with 5% $CO_2$ at 37 °C. The 96-well microtiter plates are seeded with $1.2\times10^4$ cells/100 µL/well. Plates are incubated at 37 °C in a $CO_2$ incubator overnight. The next day tested sera were heated at 56 °C for 30 min to inactivate complement, and then diluted in DMEM (supplemented with 2% FBS and 1x Penicillin/Streptomycin). Serial 2-fold dilutions of sera were carried out for the dilutions. Fifty µL of diluted sera were mixed with an equal volume of virus (100 $TCID_{50}$) and incubated at 37 °C for 1 hr. After removing the overnight culture medium, 100 µL of the sera-virus mixtures were inoculated onto a confluent monolayer of Vero-E6 cells in 96-well plates in triplicate. After incubation for 4 days at 37 °C with 5% $CO_2$, the cells were fixed with 10% formaldehyde and stained with 0.5% crystal violet staining solution at room temperature for 20 min. Individual wells were scored for CPE as having a binary outcome of 'infection" or 'no infection'. Determination of SARS-CoV-2 virus specific neutralization titer was to measure the neutralizing antibody titer against SARS-CoV-2 virus based on the principle of $VNT_{50}$ titer (≥50% reduction of virus-induced cytopathic effects). Virus neutralization titer of a serum was defined as the reciprocal of the highest serum dilution at which 50% reduction in cytopathic effects are observed and results are calculated by the method of Reed and Muench.

**Pseudovirus-neutralization assay against wild-type, BA.1 BA.2, BA.5, BQ.1.1, XBB.1.5, and CH.1.1 variants.**

Neutralizing antibody titers were measured by neutralization assay using HEK-293T-ACE2 cells challenged with SARS-CoV-2 pseudovirus variants. The study was conducted in a BSL2 lab at RNAi core, Biomedical Translation Research Center (BioTReC), Academia Sinica. Human embryonic kidney (HEK-293T/17; ATCC CRL-11268TM) cells were obtained from the American Type Culture Collection (ATCC). Cells were cultured in DMEM (Gibco) supplemented with 10% fetal bovine serum (Hyclone) and 100 U/mL of Penicillin-Streptomycin solution (Gibco), and then incubated in a humidified atmosphere with 5% $CO_2$ at 37 °C. HEK-293T-ACE2 cells were generated by transduction of VSV-G pseudotyped lentivirus carrying human ACE2 gene. To produce SARS-CoV-2 pseudoviruses, a plasmid expressing C-terminal truncated wild-type Wuhan-Hu-1 strain SARS-CoV-2 spike protein (pcDNA3.1-nCoV-SΔ18) was co-transfected into

HEK-293T/17 cells with packaging and reporter plasmids (pCMVΔ8.91, and pLAS2w.FLuc.Ppuro, respectively) (BioTReC, Academia Sinica), using TransIT-LT1 transfection reagent (Mirus Bio). Site-directed mutagenesis was used to generate the Omicron BA.1, BA.2, and BA.4/BA.5 variants by changing nucleotides from Wuhan-Hu-1 reference strain. For BA.1 variant, the mutations of spike protein are A67V, Δ69-70, T95I, G142D/Δ143-145, Δ211/L212I, ins214EPE, G339D, S371L, S373P, S375F, K417N, N440K, G446S, S477N, T478K, E484A, Q493R, G496S, Q498R, N501Y, Y505H, T547K, D614G, H655Y, N679K, P681H, N764K, D796Y, N856K, Q954H, N969K, L981F. For BA.2 variant, the mutations of spike protein are T19I, L24S, Δ25-27, G142D, V213G, G339D, S371F, S373P, S375F, T376A, D405N, R408S, K417N, N440K, S477N, T478K, E484A, Q493R, Q498R, N501Y, Y505H, D614G, H655Y, N679K, P681H, N764K, D796Y, Q954H, N969K. For BA.4/5 variant, the mutations of spike protein are T19I, L24S, Δ25-27, Δ69-70, G142D, V213G, G339D, S371F, S373P, S375F, T,376A, D405N, R408S, K417N, N440K, L452R, S477N, T478K, E484A, L486V, Q493, Q498R, N501Y, Y505H, D614G, H655Y, N679K, N764K, D796Y, N856K, and Q954H, & L969K. For BQ.1.1 variant, the mutations of spike protein are T19I, L24S, Δ25-27, Δ69-70, G142D, V213G, G339D, R346T, S371F, S373P, S375F, T376A, D405N, R408S, K417N, N440K, K444T, L452R, N460K, S477N, T478K, E484A, F486V, Q498R, N501Y, Y505H, D614G, H655Y, N679K, P681H, N764K, D796Y, Q954H, N969K. For XBB.1.5 variant, the mutations of spike protein are T19I, L24S, Δ25-27, V83A, Δ144, G142D, H146Q, Q183E, V213E, G252V, G339H, R346T, L368I, S371F, S373P, S375F, T376A, D405N, R408S, K417N, N440K, V445P, G446S, N460K, S477N, T478K, E484A, F486P, F490S, Q498R, N501Y, Y505H, D614G, H655Y, N679K, P681H, N764K, D796Y, Q954H, N969K. For CH.1.1 variant, the mutations of spike protein are T19I, L24S, Δ25-27, G142D, K147E, W152R, F157L, I210V, V213G, G257S, G339H, R346T, S371F, S373P, S375F, T376A, D405N, R408S, K417N, N440K, K444T, G446S, L452R, N460K, S477N, T478K, E484A, F486S, Q498R, N501Y, Y505H, D614G, H655Y, N679K, P681H, N764K, D796Y, Q954H, N969K. Indicated plasmids were delivered into HEK-293T/17 cells by using TransITR-LT1 transfection reagent (Mirus Bio) to produce different SARS-CoV-2 pseudoviruses. At 72 hours post-transfection, cell debris were removed by centrifugation at 4,000 xg for 10 minutes, and supernatants were collected, filtered (0.45 µm, Pall Corporation) and frozen at −80 °C until use. HEK-293-hACE2 cells (1x$10^4$ cells/well) were seeded in 96-well white isoplates and incubated for overnight. Tested sera were heated at 56°C for 30 min to inactivate complement, and diluted in medium (DMEM supplemented with 1% FBS and 100 U/ml Penicillin/Streptomycin), and then 2-fold serial dilutions were carried out for a total of 8 dilutions. The 25 µL diluted sera were mixed with an equal volume of pseudovirus (1,000 TU) and incubated at 37 °C for 1 hr before adding to the plates with cells. After 1-hr incubation, the 50 µL mixture added to the plate with cells containing with 50 µL of DMEM culture medium per well at the indicated dilution factors. On the following 16 hours incubation, the culture medium was replaced with 50 µL of fresh medium (DMEM supplemented with 10% FBS and 100 U/ml Penicillin/Streptomycin). Cells were lysed at 72 hours post-infection and relative light units (RLU) was measured by using Bright-GloTM Luciferase Assay System (Promega). The luciferase activity was detected by Tecan i-control (Infinite 500). The percentage of inhibition was calculated as the ratio of RLU reduction in the presence of diluted serum to the RLU value of virus only control and the calculation formula was shown below: (RLU Control - RLU Serum) / RLU Control. The 50% protective titer ($NT_{50}$ titer) was determined by Reed and Muench method.

**T cell responses by ELISpot.** Human peripheral blood mononuclear cells (PBMCs) were used in the detection of the T cell response. For the booster-series third-dose series extension study, ELISpot assays were performed using the human IFN-γ/IL-4 FluoroSpot$^{PLUS}$ kit (MABTECH). Aliquots of 250,000 PBMCs were plated into each well and stimulated, respectively, with 10 µg/mL (each stimulator) of $RBD\text{-}_{WT}$+Th/CTL, Th/CTL, or Th/CTL pool without UBITh1a (CoV2 peptides), and cultured in culture medium alone as negative controls for each plate for 24 hours at 37 °C with 5% $CO_2$. The analysis was conducted according to the manufacturer's instructions. Spot-forming units (SFU) per million cells was calculated by subtracting the negative control wells.

**Intracellular cytokine staining (ICS).** Intracellular cytokine staining and flow cytometry was used to evaluate $CD4^+$ and $CD8^+$ T cell responses. PBMCs were stimulated, respectively, with S1-RBD-His recombinant protein plus with Th/CTL peptide pool, Th/CTL peptide pool only, CoV2 peptides, PMA + Inonmycin (as positive controls), or cultured in culture medium alone as negative controls for 6 hours at 37°C with 5% $CO_2$. Following stimulation, cells were washed and stained with viability dye for 20 minutes at room temperature, followed by surface stain for 20 minutes at room temperature, cell fixation and permeabilization with the BD cytofix/cytoperm kit (Catalog # 554714) for 20 minutes at room temperature, and then intracellular stain for 20 minutes at room temperature. Intracellular cytokine staining of IFN-γ, IL-2 and IL-4 was used to evaluate $CD4^+$ T cell response. Intracellular cytokine staining of IFN-γ, IL-2, CD107a and Granzyme B was used to evaluate $CD8^+$ T cell responses. Upon completion of staining, cells were analyzed in a FACSCanto II flow cytometry (BD Biosciences) using BD FACSDiva software.

#### Supplemental Figures

**Figure S1. Pedigree chart of SARS-CoV-2 Omicron and distribution of major circulating Omicron subvariants**

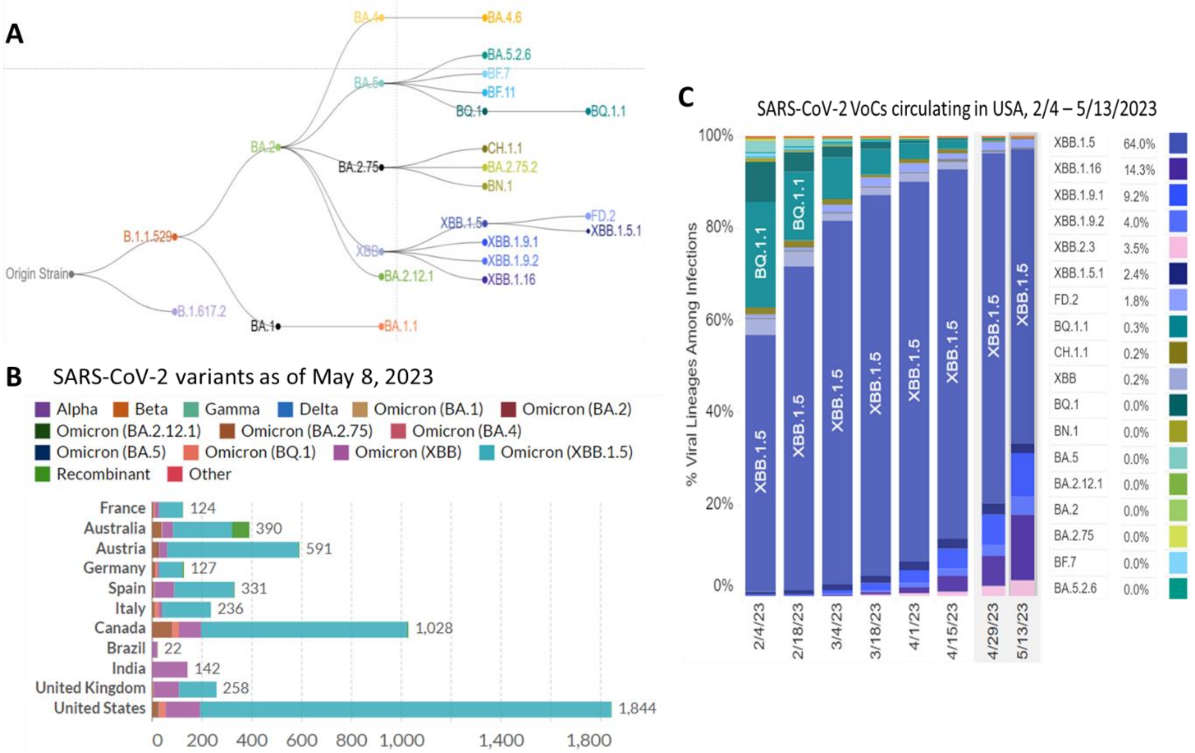

**(A)** Pedigree chart of SARS-CoV-2 subvariants derived from the original Wuhan wild-type strain. Aside from the highly virulent Delta (B.1617.2) that can infect systemically, the initial Omicron (B.1.1.529) evolves into sublineages that are heavily mutated, highly transmissible (immune-evasive) yet much less systemically infectible Omicrons. **(B)** Distributions of Omicrons worldwide showing the bulk of infection cases occurring in USA, Europe, Australia and India, with XBB.1.5 or XBB as the dominant variant. **(C)** Distributions of Omicronsin in USA USA showing XBB.1.5 is taking the lead, followed by XBB.1.16. [SARS-CoV-2 pedigree and strain distribution are *sourced from: https://covid.cdc.gov/covid-data-tracker/#variant-proportions; https://ourworldindata.org/covid-cases*]

**Figure S2. Participants of the Observational Study V-205-Q stayed on the UB-612 homologous booster (3rd dose) over time**

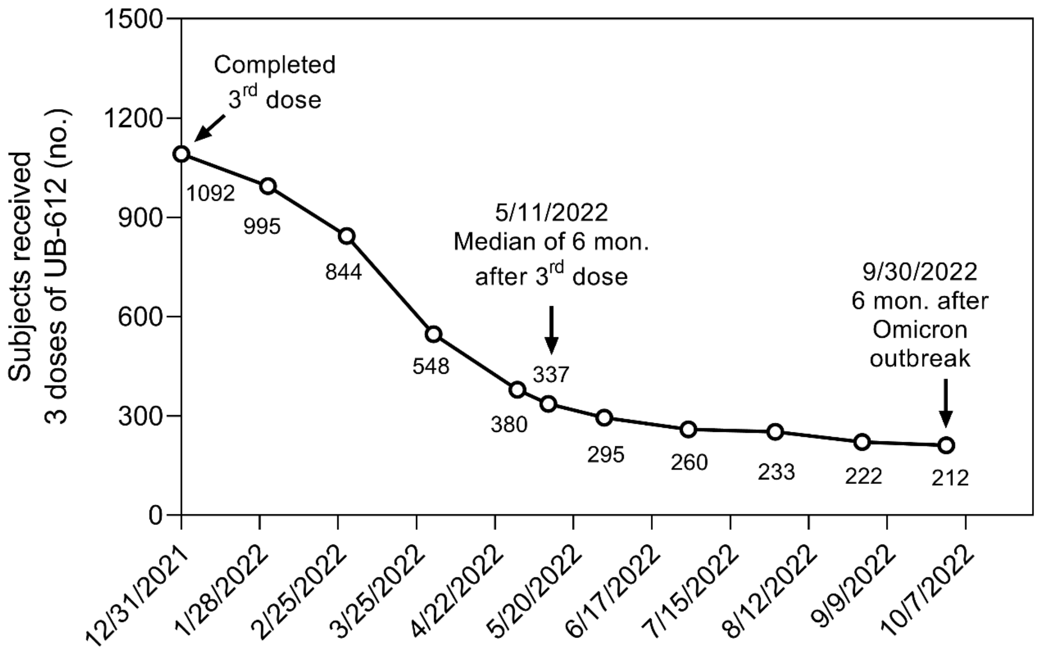

A status of non-EUA approval for UB-612 in August 2021 in Taiwan rendered low sample size for evaluation of protection effectiveness upon booster vaccination, as the bulk of the primary-2 dose vaccines were encouraged by Taiwan CDC to receive EUA-authorized vaccines for the booster 3rd dose. As such, the study participants stayed on the course of UB-612 primary and booster vaccination decreased sharply, especially after the Omicron outbreak. However, there were still 337 and 212 subjects received only 1 homologous booster without any heterologous booster on May 11, 2022 and September 30, 2022, respectively.

**Figure S3. UB-612 vaccination induced potent and durable T cell responses measured by ELISpot and ICS[a]**

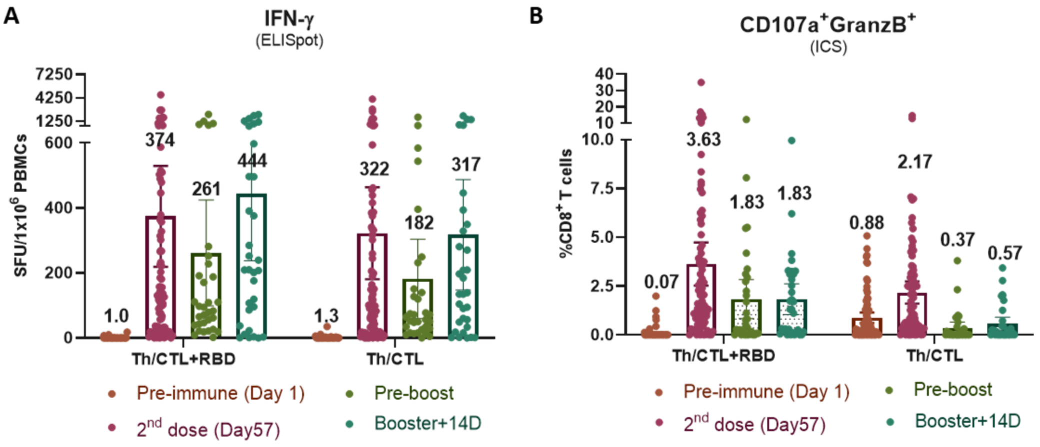

C

| Booster (SFU/$10^6$ cells) | UB-UB -UB[a] | ChA-ChA -ChA[b] | BNT-BNT -BNT[b] | BNT-BNT -BNT-BNT[c] |
|---|---|---|---|---|
| Pre-boost | 261 | 38 | 28 | 14 |
| Post-boost | 444 | 45 | 82 | 80 |

[a]UB = UB-612; [b]ChA = ChAdOx-1; [b]BNT = BNT162b2.
[b] *Lancet. 2021;398:2258-2276.* [c] *Lancet Infect Dis. 2022;21:1131-1141*

T-cell responses to stimulation by epitope peptides (RBD+Th/CTL or Th/CTL alone) were analysed with PBMCs collected from 83 vaccinees from Immunogenicity group (n = 83) on Days 57 (28 days after 2[nd] dose); and from 32 vaccinees from the Immunogenicity (n = 18) or Safety groups (n = 14) who joined the Phase-2 extension homologous booster study to evaluate the T-cell responses in PBMCs on Days 197 to 242 (pre-boosting days) and Days 211 to 256 (14 days post-booster third dose). **(A)** T-cell responses expressed as Spot-Forming Units (SFU) per 1×$10^6$ PBMCs producing IFN-γ were measured by ELISpot assay under stimulation with epitope peptides plus or without RBD. **(B)** T-cell responses expressed as %$CD8^+CD107a^+GramzB^+$ were measured by ICS under stimulation with epitope peptides plus or without RBD. **(C)** In contrast with UB-612's IFN-$\gamma^+$-T cell ELISpot under RBD+Th/CTL stimulation, the counterpart ELISpot data reported after homologous booster vaccination from ChAdOx-1 (ChA) and BNT62b2 (BNT) vaccines were laid out. BNT involved 3 and 4 homologous doses. The results indicate that the conserved Th/CTL epitope peptides in UB-612 vaccine are the principal driver of potent memory T cell immunity, which could have competitive edge over other vaccine platforms. *[[a]Panels A and B are adapted from PLOS Pathog. 2023;19(4):e1010870* https://doi.org/10.1371/journal.ppat.1010870*]*

#### Supplemental Tables

##### Table S1. Pan-Sarbecovirus vaccine UB-612 rationally designed with Th and CTL epitope eptides seuence-conserved across all sarbecoviruse strains[a]

| Sarbecoviruses WT & VoCs | M SARS-CoV2 $M_{101-156}$ (CTL epitope) | N SARS-CoV2 $N_{305-331}$ (Th/CTL epitope) | S2[b,c] SARS-CoV2 $S_{957-984}$ (Th/CTL epitope) | S2 SARS-CoV2 $S_{891-917}$ (Th epitope) | S2 SARS-CoV2 $S_{996-1028}$ (Th/CTL epitope) |
|---|---|---|---|---|---|
| **SARS-CoV-1 (WT)** | GLMWLSYFIASFRLFA RTRSMWS | AQFAPSASAFFGMSR IGMEVTPSGTWL | QALNTLVKQLSSNFGAISS VLNDILSRL | GAALQIPFAMQMAY RFNGIGVTQNVLY | LITGRLQSLQTVVTQLI RAAEIRASANLAATK |
| **SARS-CoV-2 Wuhan (Original)** | GLMWLSYFIASFRLFA RTRSMWS | AQFAPSASAFFGMSR IGMEVTPSGTWL | QALNTLVKQLSSNFGAISS VLNDILSRL | GAALQIPFAMQMAY RFNGIGVTQNVLY | LITGRLQSLQTVVTQLI RAAEIRASANLAATK |
| **Alpha, Beta, & Gamma** | GLMWLSYFIASFRLFA RTRSMWS | AQFAPSASAFFGMSR IGMEVTPSGTWL | QALNTLVKQLSSNFGAISS VLNDILSRL | GAALQIPFAMQMAY RFNGIGVTQNVLY | LITGRLQSLQTVVTQLI RAAEIRASANLAATK |
| **Delta** | GLMWLSYFIASFRLFA RTRSMWS | AQFAPSASAFFGMSR IGMEVTPSGTWL | QALNTLVKQLSSNFGAISS VLNDILSRL | GAALQIPFAMQMAY RFNGIGVTQNVLY | LITGRLQSLQTVVTQLI RAAEIRASANLAATK |
| **Omicron[c] (BA.1)** | GLMWLSYFIASFRLFA RTRSMWS | AQFAPSASAFFGMSR IGMEVTPSGTWL | QALNTLVKQLSS**K**FGAISSV LNDI**F**SRL | GAALQIPFAMQMAY RFNGIGVTQNVLY | LITGRLQSLQTVVTQLI RAAEIRASANLAATK |
| **Omicron[c] (BA.2/BA.4/ BA.5)** | GLMWLSYFIASFRLFA RTRSMWS | AQFAPSASAFFGMSR IGMEVTPSGTWL | QALNTLVKQLSS**K**FGAISSV LNDILSRL | GAALQIPFAMQMAY RFNGIGVTQNVLY | LITGRLQSLQTVVTQLI RAAEIRASANLAATK |
| **Omicron[c] (BF.7/BQ/XBB)** | GLMWLSYFIASFRLFA RTRSMWS | AQFAPSASAFFGMSR IGMEVTPSGTWL | QALNTLVKQLSS**K**FGAISSV LNDILSRL | GAALQIPFAMQMAY RFNGIGVTQNVLY | LITGRLQSLQTVVTQLI RAAEIRASANLAATK |
| **Omicron[c] (CH.1.1/CA.3.1)** | GLMWLSYFIASFRLFA RTRSMWS | AQFAPSASAFFGMSR IGMEVTPSGTWL | QALNTLVKQLSS**K**FGAISSV LNDILSRL | GAALQIPFAMQMAY RFNGIGVTQNVLY | LITGRLQSLQTVVTQLI RAAEIRASANLAATK |

[a] The presence of conserved T cell epitopes is critical for the induction of universal, cross-reactive T cell memory immunity against viral antigens. Fast responding memory T cells are pivotal in the protection against and help prevent onset of severe COVID-19. SARS-CoV-1 and SARS-CoV-2 CTL and Th epitopes, validated by HLA binding and T cell functional assays, are highly common, conserved between across Sarbecoviruse species (SARS-CoV-1 and SARS-CoV-2), with minor between-variant differences seen only at S957-984. The Wuhan wild-type peptides (M, N and S2x3) are employed for precision-design of UB-612 vaccine against COVID-19. Identification of T cell epitopes on SARS-CoV-1 (2003), determined using HLA-binding assays, were used to determine corresponding T cell epitopes in SARS-CoV-2 (2019) by sequence alignment (*Patent WO2021/168305A1, 2020, CY Wang et al.).* In this Table 1, the sequence-conserved epitopes have been updated to cover subvariants related to BF.7/BQ/XBB and CH.1/CA.3.1, in addition to the earliest SARS-CoV-1. To justify that the targeted epitopes are essentially universal and UB-612 represents a genre of pan-Sarbecovirus vaccine.

[b] Except for N969K (on BA.1 through CH.1.1) and L981F (on BA.1) within S957-984 peptide on the S2 spike protein, none of the other four designer epitope peptides for UB-612 vaccine has an aa-residue that overlaps with the reported mutation sites on Spike, M, and N proteins protein (Supplemental Table 5).

[c] At S957-984, there are minor sequence differences between Omicron BA.1 and BA.2/BA.4/BA.5, marked in boldface and underscored. BF.7, BQ, XBB, CH.1.1, and CA.1.3 have the same mutation as BA2/BA.4/BA.5 at N969K.

**Table S2. Omicron-induced COVID-19 outbreak events in Taiwan**

| Date | Outbreak events[a] |
|---|---|
| 01-09-2022 | Median day of 7 days average >50 confirmed cases/day |
| 03-21-2022 | Median day of 7 days average >100 confirmed cases/day |
| 04-09-2022 | Median day of 7 days average >500 confirmed cases/day |
| 04-14-2022 | Median day of 7 days average >1,000 confirmed cases/day |
| 04-28-2022 | Median day of 7 days average >10,000 confirmed cases/day |
| 05-05-2022 | Accumulated 232,402 (1.0 %) confirmed cases |
| 05-11-2022 | Accumulated 505,455 (2.2 %) confirmed cases |
| 05-21-2022 | Accumulated 1,240,897 (5.4 %) confirmed cases |
| 06-04-2022 | Accumulated 2,342,794 (10.1 %) confirmed cases in Taiwan |
| 09-30-2022 | Accumulated 6,461,400 (27.9 %) confirmed cases in Taiwan |

[a] The Omicron outbreak is defined as the median day of 7-days average >100 confirmed cases/day. Shown in the table are the early phase of Omicron outbreak in March and April 2022 (7-days average confirmed cases/day) and the log phase (accumulated confirmed cases) ranging from May through September 2022.

**Table S3. Post-booster live virus-neutralizing antibody titers ($VNT_{50}$) against Wuhan wild-type (WT) and Omicron BA.1, BA.2 and BA.5 by UB-612 and other vaccine platforms**

| Vaccine | WT (booster) | BA.1 (booster) | BA.2 (booster) | BA.5 (booster) |
|---|---|---|---|---|
| UB-612 | 2424 | 359 | 325 | 123 |
| mRNA-1273[a] | 1,659 | 81.0 | NA | NA |
| BNT162b2[a] | 640 | 46.2 | NA | NA |
| BNT162b2[b] | 673 | 106 | NA | NA |
| AZD1222[c] | 723 | 57 | NA | NA |

UB-612: live virus-neutralizing data after booster vaccination from the Figure 4 of this manuscript. NA: not available.

[a]*Cell Rep Med.* 2022:100679. https://doi.org/10.1016/j.xcrm.2022.100679
[b]*Science.* 2022;375:678-680. [c]*Cell*. 2022;185:467-484.

**Table S4. UB-612 can serve as a competent heterologous booster enhancing viral-neutralizing titer and seroconversion rate***

Fold increase (FI) of pseudovirus-neutralizing titer upon booster

| Heterologous (Ht) | Homologous (Hm) | WT (FI by Ht/Hm) | BA.5 (FI by Ht/Hm) |
|---|---|---|---|
| BNT-BNT-**UB** | BNT-BNT-BNT | 1.04 | 1.11 |
| ChA-ChA-**UB** | ChA-ChA-ChA | 1.92[a] | 2.85[a] |
| BIBP-BIBP-**UB** | BIBP-BIBP-BIBP | 5.77[a] | 5.93[a] |

WT: Wuhan wild type. BA.5: Omicron BA.5 variant. BNT: BNT162b2 vaccine. ChA: ChAdOx1-S vaccine
BIBP: Sinovac inactivated vaccine. UB: UB-612 vaccine. [a] P<0.0001

Fold increase (FI) of seroconversion rate upon booster

| Heterologous (Ht) | Homologous (Hm) | WT (FI by Ht/Hm) | BA.5 (FI by Ht/Hm) |
|---|---|---|---|
| BNT-BNT-**UB** | BNT-BNT-BNT | ~1.00 | ~1.00 |
| ChA-ChA-**UB** | ChA-ChA-ChA | 1.9[b] | 2.0[c] |
| BIBP-BIBP-**UB** | BIBP-BIBP-BIBP | 8.3[c] | 5.8[c] |

WT: Wuhan wild type. BA.5: Omicron BA.5 variant. BNT: BNT162b2 vaccine. ChA: ChAdOx1-S vaccine
BIBP: Sinovac inactivated vaccine. UB: UB-612 vaccine. [b] P<0.0009; [c] P<0.0001

* https://www.globenewswire.com/news-release/2022/12/02/2566668/0/en/Vaxxinity-Announces-Positive-Topline-Pivotal-Phase-3-COVID-19-Booster-Data-for-UB-612.html

#### Table S5. Mutation sites on SARS-CoV-2 spike (S), envelope (E), membrane (M), and nucleocapsid (N) proteins in Omicrons

| VoC | Spike (S1-RBD residues at 319-541) | E | M | N |
|---|---|---|---|---|
| Omicron (BA. 4) | T19I, L24S, Δ25-27, Δ69-70, G142D, V213G, G339D, S371F, S373P, S375F, T,376A, D405N, R408S, K417N, N440K, L452R, S477N, T478K, E484A, L486V, Q493, Q498R, N501Y, Y505H, D614G, H655Y, N679K, N764K, D796Y, N856K, and Q954H, & L969K | T9I | Q19E & A63T | P13L, Δ31-33, P151S, R203K, G204R, & S413R |
| Omicron (BA.5) | T19I, L24S, Δ25-27, Δ69-70, G142D, V213G, G339D, S371F, S373P, S375F, T,376A, D405N, R408S, K417N, N440K, L452R, S477N, T478K, E484A, L486V, Q493, Q498R, N501Y, Y505H, D614G, H655Y, N679K, N764K, D796Y, N856K, and Q954H, & L969K | T9I | D3N, Q19E, & A63T | P13L, Δ31-33, R203K, G204R, & S413R |
| Omicron (BQ.1) | T19I, L24S, Δ25-27, Δ69-70, G142D, V213G, G339D, S371F, S373P, S375F, T376A, D405N, R408S, K417N, N440K, K444T, L452R, N460K, S477N, T478K, E484A, F486V, Q498R, N501Y, Y505H, D614G, H655Y, N679K, P681H, N764K, D796Y, Q954H, N969K | T9I | D3N, Q19E, & A63T | P13L, Δ31-33, E136D,R203K, G204R, & S413R |
| Omicron (BQ.1.1) | T19I, L24S, Δ25-27, Δ69-70, G142D, V213G, G339D, R346T, S371F, S373P, S375F, T376A, D405N, R408S, K417N, N440K, K444T, L452R, N460K, S477N, T478K, E484A, F486V, Q498R, N501Y, Y505H, D614G, H655Y, N679K, P681H, N764K, D796Y, Q954H, N969K | T9I | D3N, Q19E, & A63T | P13L, Δ31-33, E136D,R203K, G204R, & S413R |
| Omicron (XBB.1) | T19I, L24S, Δ25-27, V83A, Δ144, G142D, H146Q, Q183E, V213E, G252V, G339H, R346T, L368I, S371F, S373P, S375F, T376A, D405N, R408S, K417N, N440K, V445P, G446S, N460K, S477N, T478K, E484A, F486S, F490S, Q498R, N501Y, Y505H, D614G, H655Y, N679K, P681H, N764K, D796Y, Q954H, N969K | T9I, T11A | Q19E, A63T | P13L, Δ31-33, R203K, G204R, & S413R |
| Omicron (XBB.1.5) | T19I, L24S, Δ25-27, V83A, Δ144, G142D, H146Q, Q183E, V213E, G252V, G339H, R346T, L368I, S371F, S373P, S375F, T376A, D405N, R408S, K417N, N440K, V445P, G446S, N460K, S477N, T478K, E484A, F486P, F490S, Q498R, N501Y, Y505H, D614G, H655Y, N679K, P681H, N764K, D796Y, Q954H, N969K | T9I, T11A | Q19E, A63T | P13L, Δ31-33, R203K, G204R, & S413R |
| Omicron (CH.1.1) | T19I, L24S, Δ25-27, G142D, K147E, W152R, F157L, I210V, V213G, G257S, G339H, R346T, S371F, S373P, S375F, T376A, D405N, R408S, K417N, N440K, K444T, G446S, L452R, N460K, S477N, T478K, E484A, F486S, Q498R, N501Y, Y505H, D614G, H655Y, N679K, P681H, N764K, D796Y, Q954H, N969K | T9I, T11A | Q19E, A63T | P13L, Δ31-33, R203K, G204R, & S413R |

*https://viralzone.expasy.org/955/* ; *https://outbreak.info/situation-reports#voc*

### Appendix 1. Phase-2 study V-205-Q protocol

### CLINICAL STUDY PROTOCOL

#### *A Questionnaire-based, Retrospective, Phase II Extension Study to Investigate the COVID-19 Breakthrough Infection after 2 doses of UB-612 Vaccination in Adolescent, Younger and Elderly Adult Volunteers*

| | |
|---|---|
| **Protocol Number:** | V-205-Q |
| **Investigational Product:** | UB-612 |
| **Phase:** | Exploratory study |
| **Sponsor:** | 聯亞生技開發股份有限公司<br>United Biomedical, Inc., Asia (UBI Asia),<br>Hsinchu County, Taiwan |
| **Protocol Date:** | November 22, 2022 |
| **Protocol Version:** | 2.0 |

CONFIDENTIAL
**This protocol may not be reproduced or communicated to a third party without the written permission of United Biomedical, Inc., Asia**

### 1. PROTOCOL APPROVAL SIGNATURES

**Protocol Title:** A Questionnaire-based, Retrospective, Phase II Extension Study to Investigate the COVID-19 Breakthrough Infection after 2 doses of UB-612 Vaccination in Adolescent, Younger and Elderly Adult Volunteers

**Protocol Number:** V-205-Q

This study will be conducted in compliance with the clinical study protocol (and amendments), International Council for Harmonisation (ICH) guidelines for current Good Clinical Practice (cGCP) and applicable regulatory requirements.

**Sponsor Signatory**

Chang Yi Wang
United Biomedical, Inc., Asia (UBI Asia),
No. 45, Guangfu N. Rd., Hukou Township,
Hsinchu County 303, Taiwan (R.O.C.)


Signature

Nov 23rd, 2022
Date

#### 2. SYNOPSIS

**Protocol Number:**

V-205-Q

**Title:**

A Questionnaire-based, Retrospective, Phase II Extension Study to Investigate the COVID-19 breakthrough infection after 2 doses of UB-612 Vaccination in Adolescent, Younger and Elderly Adult Volunteers

**Investigational Product**:

UB-612 vaccine

**Study Design:**

This is an extension and retrospective questionnaire to survey the COVID-19 breakthrough infection in volunteers completed V-205 phase II study. Study population consists of adult and adolescent subjects who had ever received at least two UB-612 vaccinations (the 1$^{st}$ dose and the 2$^{nd}$ dose) in the V-205 phase II study. Eligible subjects will be contacted by telephone to obtain the consent to questionnaire survey. The questionnaire contains those participants' SARS-COV-2 infection history and the corresponding disease severity after UB-612 2-dose primary vaccination series.

**Number of Subjects:**

It expects to recruit up to 3,300 adult and 354 adolescent participants.

**Study Population:**

<u>Inclusion criteria</u>

To be eligible for study entry, subjects must satisfy all of the following inclusion criteria:

1. Male or female who previously participated in and completed at least 2-dose primary vaccination (1$^{st}$ and 2$^{nd}$ dose) in the V-205 phase II Study.
2. Participants or the participants' legal representatives must agree to join questionnaire survey.
3. Participants or the participants' legal representative must understand and agree to complete the e-questionnaire.

**Objectives and Endpoint(s):**

| **Objectives** | **Endpoints** | **Groups** |
|---|---|---|
| Primary | | |
| To survey the disease severity of COVID-19 cases which occurred in participants who completed the 2-dose primary vaccination series | The number and percentage of COVID-19 cases with the occurrence of the severity:<br>• Asymptomatic infection<br>• Mild: had symptoms without hospitalization<br>• Moderate: hospitalized due to infection<br>• Severe: hospitalized in intensive care unit due to infection | • Group 1<br>• Group 2<br>• Group 3<br>• Group 4<br>• Overall |
| Secondary | | |
| To obtain the numbers of COVID-19 cases which occurred in participants who completed the 2-dose primary vaccination series | The number of participants with the occurrence of:<br>• COVID-19 cases confirmed by receiving "COVID-19 Designated Residence Isolation (Home Isolation) Notice and Right to Petition for Habeas Corpus Relief". | • Group 1<br>• Group 2<br>• Group 3<br>• Group 4<br>• Overall |
| To describe risk factors for developing COVID-19 | The number and proportion of COVID-19 based on the following risk factors:<br>• Age<br>• Sex<br>• Comorbidity<br>• Obesity (BMI≥30 kg/m$^2$) | • Group 1<br>• Group 2<br>• Group 3<br>• Group 4<br>• Overall |
| To describe risk factors for developing moderate and severe COVID-19 illness | The number and proportion of moderate and severe COVID-19 illness based on the following risk factors:<br>• Age<br>• Sex<br>• Comorbidity<br>• Obesity (BMI≥30 kg/m$^2$) | • Group 1<br>• Group 2<br>• Group 3<br>• Group 4<br>• Overall |
| To investigate the time to first confirmed SARS-CoV-2 infection after 2-dose primary vaccination series | The time to first confirmed SARS-CoV-2 infection after 2-dose primary vaccination series and before other vaccination<br>• The date of first confirmed SARS-CoV-2 infection is defined as the 1st day of | • Group 1<br>• Group 2<br>• Group 3<br>• Group 4<br>• Group 1/2 |

| | | |
|---|---|---|
| | residence isolation recorded in "COVID-19 Designated Residence Isolation (Home Isolation) Notice and Right to Petition for Habeas Corpus Relief" | • Group 3/4 |
| Exploratory | | |
| To evaluate incidence rate of COVID-19 cases which occurred in participants who completed the 2-dose primary vaccination series | The incidence rate of participants presented as a percentage with the occurrence of:<br>• COVID-19 cases confirmed by receiving "COVID-19 Designated Residence Isolation (Home Isolation) Notice and Right to Petition for Habeas Corpus Relief". | • Group 1<br>• Group 2<br>• Group 3<br>• Group 4<br>• Overall |
| To estimate the influence of potential environmental factors in participants who completed the 2-dose primary vaccination series | The number and proportion of participants based on the following potential environmental factors:<br>• The definition of co-residents: Person(s) live(s) at the same address with the participant for more than one day<br>• Number and proportion of participants without COVID-19, living with co-residents who infected with SARS-COV-2.<br>• Number and proportion of participants' infectious period which overlapped the infectious period of co-residents within 14 days. | • Group 1<br>• Group 2<br>• Group 3<br>• Group 4<br>• Overall |
| To evaluate influence factors for the time to first confirmed SARS-CoV-2 infection after 2-dose primary vaccination series | The time to first confirmed SARS-CoV-2 infection after 2-dose primary vaccination series affected by the following factors:<br>• Age<br>• Sex<br>• Comorbidity<br>• Obesity (BMI≥30 kg/m$^2$) | • Group 1<br>• Group 2<br>• Group 3<br>• Group 4<br>• Overall |
| Compared with external information, incidence rate of COVID-19 | The comparison between the external information described in section 8.4 of | • Group 1<br>• Group 2<br>• Group 3 |

| | | |
|---|---|---|
| | protocol and the data collected in this study in terms of the following items:<br>• Incidence rate of COVID-19<br>• The rate of breakthrough infections after primary vaccination series<br>• The rate of breakthrough infections after the booster vaccination | • Group 4 |

Definition of study groups is as below.

| Group | Description |
|---|---|
| Group 1 | Participants who received two doses of UB-612 and did not receive COVID-19 vaccines other than UB-612 before study enrolment |
| Group 2 | Participants who received two doses of UB-612 and then received COVID-19 vaccine(s) other than UB-612 before study enrolment |
| Group 3 | Participants who received three doses of UB-612 and did not receive COVID-19 vaccines other than UB-612 before study enrolment |
| Group 4 | Participants who received three doses of UB-612 and then received COVID-19 vaccine(s) other than UB-612 before study enrolment |
| Overall | Pooling Group 1 to Group 4 |

**<u>Statistical Analysis</u>**

All the data summary is based on subjects who have the valid questionnaire record. The available data will be displayed via the descriptive statistics no statistical testing is adopted. For continuous variables, the number, mean, standard deviation, median, minimum, and maximum values will be presented. For categorical variables, the numbers and percentages of subjects in each class will be presented. All confidence intervals, if provided, will be the 95% confidence intervals. For time to event data, the Kaplan-Meier approach will be used in analysis with Kaplan-Meier plot. All available data from the valid questionnaire will be displayed and utilized in data analysis. No imputation will be considered for the missing observations from the subjects reported e-questionnaire.

#### 4. LIST OF ABBREVIATIONS

| | |
|---|---|
| ACE2 | Angiotensin-converting enzyme 2 |
| ADE | Antibody dependent enhancement (of viral replication) |
| AE | adverse event |
| BMI | body mass index |
| CECC | Central Epidemic Command Center |
| CDC | Centers for Disease Control and Prevention |
| cGCP | current good clinical practice |
| COVID-19 | corona virus disease 2019 |
| CRF | case report form |
| CTL | cytotoxic T Lymphocyte |
| EUA | Emergency Use Authorization |
| FDA | Food and Drug Administration |
| GMFI | geometric mean fold increase |
| GMT | geometric mean titer |
| ICF | informed consent form |
| ICH | International Council for Harmonisation |
| ICTV | International Committee on Taxonomy of Viruses |
| IEC | independent ethics committee |
| IRB | institutional review board |
| MHC | major histocompatibility complex |
| ORF | open reading frame |
| RBD | receptor binding domain |
| RNA | ribonucleic acid |
| RT-PCR | reverse transcriptase- polymerase chain reaction |
| SCR | seroconversion rate |
| SAE | serious adverse event |
| SRsFc | S1-RBD-sFc |
| SUSAR | suspected unexpected serious adverse reaction |
| UB-612 | United Biomedical, Inc.'s COVID-19 vaccine |
| VAERD | vaccine-associated enhanced respiratory disease |

### 5. INTRODUCTION

#### 5.1. Study Rationale

Administration 100 µg of UB-612 vaccine showed safe and tolerable profile with adequate immune response in phase I (V-122, V-123) and II (V-205) study. During early stage of the UB-612 study, the low SARS-CoV-2 infection and death rate in Taiwan were encountered due to the "zero-COVID" policy of strict border controls, meticulous tracking and tracing, and universal use of face masks. However, the omicron outbreak in early 2022 forced Taiwan to move from preventing infections to co-existing with the virus. Under the rapidly changing circumstances of COVID-19 pandemic due to higher omicron infection rate, a study for COVID-19 breakthrough cases will be valuable to investigate the effectiveness of UB-612.

#### 5.2. Background

In December 2019, a cluster of patients with pneumonia surfaced in Wuhan, China. The culprit was quickly identified as a beta-coronavirus that has never been reported before, and the disease was named by WHO as Corona Virus Disease 2019 (COVID-19) and the virus that causes it by the International Committee on Taxonomy of Viruses (ICTV) as SARS-CoV-2 [1, 2]. As of January 7, 2020, a global outbreak has caused 87,200,000 confirmed cases in more than 220 countries or territories, with 1,880,000 deaths, making the SARS-CoV-2 pandemic a general public health event that has stirred up worldwide attention. The B.1.1.529 (omicron) variant of SARS-CoV-2 has rapidly spread as the dominant variant in many countries since its first detection in early November 2021. Currently, the epidemic is still spreading. The data on the real-world effectiveness of the vaccine against omicron infection and COVID-19-related hospitalization rates in adults, the elderly, and adolescents are unclear.

SARS-CoV-2 is a positive-strand RNA virus that belongs to the group of Beta coronaviruses. The genome of SARS-CoV-2 is approximately 29,700 nucleotides long and shares 79.5% sequence identity with SARS-CoV [3]. The long ORF1ab polyprotein at 5' end of the genome encodes 15 or 16 non-structural proteins, and the 3' end encodes 4 major structural proteins, including the spike (S) protein, nucleocapsid (N) protein, membrane (M) protein, and the envelope (E) protein [4]. SARS-CoV-2 interacts with the receptor angiotensin converting enzyme 2 (ACE2) on host cells via receptor binding domain (RBD) of its S protein for viral entry and subsequent pathogenesis [5], resulting in severe respiratory illness with symptoms of fever, cough, and shortness of breath, and even death in severe cases [6].

Vaccines are the most effective and economical means to prevent and control infectious diseases [7]. The development of an effective vaccine against SARS-Co-2 infection is urgently required. Currently, more than 200 pharmaceutical companies and academic institutions worldwide have launched their programs on vaccine development against SARS-CoV-2. There are several different types of vaccines under development; one of them is subunit vaccine. Subunit vaccines include one or more antigens with strong immunogenicity capable of efficiently stimulating the host immune system. In general, this type of vaccine is safer and easier to produce, but often requires the addition of adjuvants to elicit a strong protective immune response. So far, several institutions have initiated programs on the SARS-CoV-2 subunit vaccine, and almost all of them use the S protein as antigens. For example, the University of Queensland is developing a subunit vaccine based on the "molecular clamp" technology [8]. Clover Biopharmaceuticals Inc. revealed that they are developing a vaccine candidate against SARS-CoV-2 using the "Trimer-Tag" technology [9], and the trimeric S protein subunit vaccine candidate was produced via a mammalian cell expression system. Novavax, Inc. announced that they had produced multiple nanoparticle vaccine candidates based on S protein, and after assessing efficacy in animal models to identify an optimal vaccine candidate, began phase I clinical testing in May, 2020. Besides, Johnson & Johnson, Pasteur Institute, Sanofi Pasteru, GSK, and Chongqing Zhifei Biological Products Co., Ltd. also started subunit vaccine development against SARS-CoV-2.

Safety is the most important issue that should be taken into consideration during drug and vaccine development, and some scientists urge that we should not rush to deploy COVID-19 vaccines and drugs without sufficient safety guarantees [10]. There have been concerns regarding vaccine-associated enhanced respiratory disease (VAERD) by certain candidate COVID-19 vaccine approaches, via antibody-dependent enhancement (ADE) or development of Th2 immunopathology [11]. Grifoni et al. [12] revealed predominant Th1 responses in convalescing COVID-19 cases, with little to no Th2 cytokines. Clearly more studies are required, but the data Grifoni et al. shown appear to predominantly represent a classic Th1 response to SARS-CoV-2.

##### 5.3. UB-612 COVID-19 Vaccine

United Biomedical, Inc. (UBI) has developed a vaccine candidate against SARS-CoV-2 that is designed to activate both humoral and cellular responses. For SARS-CoV-2 immunogens, UB-612 includes a designer S1-RBD-sFc (SRsFc) fusion protein formulated with designer Th and CTL epitope peptides selected from immunodominant M, S2 and N regions known to bind to human MHC I and II. This mixture of designer Th/CTL peptides is designed to elicit T cell activation, memory recall and effector functions similar to that of natural COVID-19. The S1-RBD-sFc fusion protein incorporates both linear and conformation epitopes and induces high affinity antibodies to the RBD of SARS-CoV-2. The immunogen components are formulated with an oligonucleotide containing unmethylated CpG motifs and Adju-Phos® adjuvants, which promotes the activation of antigen-presenting cells pathways to induce an optimal immunogenicity profile and achieve the prevention purpose.

In summary, UB-612 vaccine design composition (S1-RBD-sFc+Th and CTL peptides + CpG, formulated with Adju-Phos®) is expected not only to be safe and induce high titers of neutralizing antibodies, but also to provide T cell memory for a long lasting protection against SARS-COV-2 across all human subjects irrespective of age, sex and ethnicities.

##### 5.4. Phase II study summary [13]

UB-612 vaccine phase II study was began in November, 2020 which was a observer-blind, multiple-centre, randomized, placebo-controlled study to evaluate the immunogenicity, safety, tolerability and lot consistency of study vaccine in adolescent, younger and elderly adults. This study was carried out in two groups, including core group for EUA application and supplementary group for broader indication in adolescents. All subjects were randomly allocated to receive 2 doses of 100 µg vaccine or placebo, spaced 28 days apart, in a 6:1 ratio. When subjects were unblinded after Day 197, the subjects in vaccine group could receive 3$^{rd}$ dose of vaccine. In the co-primary immunogenicity endpoints, geometric mean titer (GMT) and seroconversion rate (SCR) of SARS-CoV-2 neutralizing antibody on 28 days after 2$^{nd}$ vaccination (Day 57) were observed. Local reactions and systemic events, unsolicited AEs, and SAEs were listed in the primary safety endpoints.

A total number of 4625 subjects were enrolled, in which 4296 subjects were randomly assigned in UB-612 vaccine group (3681 subjects) and placebo group (615 subjects). In Core Group (younger and elderly adults), a total number of 4142 adult subjects were enrolled, in which 3877 adult subjects were randomly assigned in UB-612 vaccine group (3323 adults) and placebo group (554 adults). There were 3875 adult subjects received the first vaccination (UB-612: 3321 adults, placebo: 554 adults) and 3844 adult subjects received the second vaccination (UB-612 vaccine: 3300 adults, placebo: 544 adults). There were 1478 adult subjects in UB-612 vaccine group received their third dose of UB-612 vaccine. In Supplementary Group (adolescents), a total number of 483 adolescents were enrolled, in which 419 adolescents were randomly assigned in UB-612 vaccine group (358 adolescents) and placebo group (61 adolescents). There were 417 adolescents received the first vaccination (UB-612 vaccine: 357 adolescents, placebo: 60 adolescents) and 414 adolescents received the second vaccination (UB-612 vaccine: 354 adolescents, placebo: 60 adolescents). No adolescents received the third dose of UB-612 vaccine.

In Core Group, the GMT of neutralizing antibody responses was 2.58 at baseline and increased to 87.04 at Day 57. At Day 197, the GMT decreased to 33.16. For the subjects received placebo, the GMT of neutralizing antibody responses was 2.60 at baseline and 2.71 at Day 57. At Day 197, the GMT was 3.23. Significant difference between vaccine group and placebo group in the GMTs of neutralizing antibody at Day 57 and Day 197 were noted with P-values < 0.0001. After Day 197 procedure completed, 1478 adult subjects in vaccine group went back to site to receive booster vaccination at Visit 6 (Day 197~Day 242). For the subjects received the third dose in the evaluable immunogenicity population, the GMT before vaccination at Visit 6 was 43.26, and the GMT at 14 days after Visit 6 increased to 744.15. In Supplementary Group, the GMT of neutralizing antibody increased from 2.62 at baseline to 114.20 at Day 57. At Day 197, the GMT decreased to 58.39. For adolescent subjects received placebo, the GMTs of neutralizing antibody were 2.89 at baseline, and 2.50 at Day 197. The GMTs of neutralizing antibody at Day 57 and at Day 197 were significantly different between UB-612 vaccine and placebo groups (P-values < 0.0001).

In Core Group, seroconversion rate (SCR) of neutralizing antibody was 95.08% at Day 57. In placebo group, SCR was 1.42% at Day 57. Significant difference in SCR between UB-612 vaccine and placebo groups was demonstrated with P-value < 0.0001. In Supplementary Group, SCR of neutralizing antibody was 97.90% at Day 57. In placebo group, SCR was 0% at Day 57. Significant difference in SCR between UB-612 vaccine and placebo groups was demonstrated with P-value < 0.0001.

In Core Group, the GMFIs of neutralizing antibody in UB-612 vaccine and placebo groups were 33.797 and 1.045 at Day 57, respectively. At Day 197, the GMFIs of neutralizing antibody in UB-612 vaccine and placebo groups were 12.389 and 1.226, respectively. Significant difference in GMFI between UB-612 vaccine and placebo groups at Day 57 and Day 197 were noted (P-values < 0.0001). For the subjects received the third dose of UB-612 vaccine in the evaluable immunogenicity population, the GMFI before the third vaccination at Visit 6 was 17.304 and the GMFI at 14 days after Visit 6 was 297.661.

After primary any vaccination (1$^{st}$ dose and 2$^{nd}$ dose) in Core group, 81.06% subjects in the UB-612 vaccine and 62.21% of subjects in placebo group had reported at least one solicited adverse event within 14 days. The local reactions within 7 days after primary any vaccination in the vaccine group and the placebo group were reported in 71.98% subjects and 28.93% subjects, respectively. In the UB-612 vaccine group, the most commonly reported local reactions were pain at injection site (67.75%) and swelling (35.96%). Systemic events within 7 days after primary any vaccination were reported in 63.32% subjects of the UB-612 vaccine group and 57.14% subjects of placebo group. In the UB-

612 vaccine group, the most commonly reported systemic events were fatigue/tiredness (44.89%), muscle pain (40.54%) and headache (23.65%). After the third dose of UB-612, 73.02% subjects had reported at least one solicited adverse event within 14 days. Systemic events within 7 days after the third dose was reported in 53.37% adult subjects. The most commonly reported systemic events were fatigue/tiredness (37.19%), muscle pain (34.16%) and headache (19.15%). Unsolicited adverse events were recorded in 52.18% subjects in vaccine group and 39.71% subjects in placebo group, respectively.

In Supplementary Group, after primary any vaccination, 91.04% adolescent subjects in UB-612 vaccine and 66.67% of adolescent subjects in placebo group had reported at least one solicited adverse event within 14 days. The local reactions within 7 days after primary any vaccination were reported in 84.31% and 31.67% adolescent subjects in vaccine group and the placebo group, respectively. In vaccine group, the most commonly reported local reactions were pain at injection site (78.15%) and swelling (31.65%). Systemic events within 7 days after any vaccination were reported in 69.75% adolescents of UB-612 vaccine group and 65.00% subjects of placebo group. In UB-612 vaccine group, the most commonly reported systemic events were muscle pain (47.62%), fatigue/tiredness (45.94%) and headache (26.33%). Unsolicited adverse events were recorded in 36.69% adolescent subjects in UB-612 vaccine group and 28.33% adolescent subjects in placebo group.

There were 76 subjects (2.29%) in vaccine group and 8 adult subjects (1.44%) in placebo group reported the serious unsolicited adverse events (SAEs). One death in UB-612 group reported due to SAEs of cardiac tamponade and haemothorax which was unrelated to UB-612 vaccine. Two adult subjects who had potential COVID-19 illness and confirmed as SARS-COV-2 infection with the positive RT-PCR results. One subject was in placebo group; another one was 44 year-old male subjects in UB-612 group had COVID-19 reported at 33 days after 2$^{nd}$ vaccination, and he was noticed to discharge due to 10 days after symptoms onset and the CT value over 30.

In conclusion, administration of UB-612 100 μg vaccine showed high neutralizing antibody responses in the vaccine groups with comparable safety profile.

##### 5.5. Risk/Benefit Assessment

Administration 100 μg of UB-612 vaccine were safe and tolerable with adequate immune response in primary series two doses and the following one booster dose in phase I (V-122 study), I extension (V-123 study), and II study (V-205 study).

The Central Epidemic Command Center(CECC) in Taiwan has advocated the fourth COVID vaccine dose (the second booster shot) to residents of long-term care facilities, medical workers, individuals aged 18 and above, workers at airports and other ports of entry, and people aged 50 and over in late July, 2022. Because Taiwan government strongly encouraged people to receive official approved COVID-19 vaccines, most of subjects in UB-612 trails may be forced to receive other brand of COVID-19 vaccine after completed V-205 study. In addition, heterogeneous vaccinations against COVID-19 for both primary series and boosters were acceptable in Taiwan's policy which also increase the complexity of vaccine research. However, the breakthrough rate after 2-dose vaccination from 2022-01-02 to 2022-05-14 was 3.36%, and the rate after booster vaccination was 3.03% according to Taiwan CDC data [14].

In order to preliminary clarify the breakthrough infection after UB-612 primary vaccination series, we aim to observe the numbers and percentage of breakthrough cases with corresponding disease severity by e-questionnaire in this phase II extension study.

#### 6. STUDY OBJECTIVES & ENDPOINT

| **Objectives** | **Endpoints** | **Groups** |
|---|---|---|
| Primary | | |
| To survey the disease severity of COVID-19 cases which occurred in participants who completed the 2-dose primary vaccination series | The number and percentage of COVID-19 cases with the occurrence of the severity:<br>• Asymptomatic infection<br>• Mild: had symptoms without hospitalization<br>• Moderate: hospitalized due to infection<br>• Severe: hospitalized in intensive care unit due to infection | • Group 1<br>• Group 2<br>• Group 3<br>• Group 4<br>• Overall |
| Secondary | | |
| To obtain the numbers of COVID-19 cases which occurred in participants who completed the 2-dose primary vaccination series | The number of participants with the occurrence of:<br>• COVID-19 cases confirmed by receiving "COVID-19 Designated Residence Isolation (Home Isolation) Notice and Right to Petition for Habeas Corpus Relief". | • Group 1<br>• Group 2<br>• Group 3<br>• Group 4<br>• Overall |
| To describe risk factors for developing COVID-19 | The number and proportion of COVID-19 based on the following risk factors:<br>• Age<br>• Sex<br>• Comorbidity<br>• Obesity (BMI≥30 kg/m$^2$) | • Group 1<br>• Group 2<br>• Group 3<br>• Group 4<br>• Overall |
| To describe risk factors for developing moderate and severe COVID-19 illness | The number and proportion of moderate and severe COVID-19 illness based on the following risk factors:<br>• Age<br>• Sex<br>• Comorbidity<br>• Obesity (BMI≥30 kg/m$^2$) | • Group 1<br>• Group 2<br>• Group 3<br>• Group 4<br>• Overall |
| To investigate the time to first confirmed SARS-COV-2 infection after 2- | The time to first confirmed SARS-COV-2 infection after 2-dose primary | • Group 1<br>• Group 2<br>• Group 3 |

| | | |
|---|---|---|
| dose primary vaccination series | vaccination series and before other vaccination<br>• The date of first confirmed SARS-COV-2 infection is defined as the 1st day of residence isolation recorded in "COVID-19 Designated Residence Isolation (Home Isolation) Notice and Right to Petition for Habeas Corpus Relief" | • Group 4<br>• Group 1/2<br>• Group 3/4 |
| Exploratory | | |
| To evaluate incidence rate of COVID-19 cases which occurred in participants who completed the 2-dose primary vaccination series | The incidence rate of participants presented as a percentage with the occurrence of:<br>• COVID-19 cases confirmed by receiving "COVID-19 Designated Residence Isolation (Home Isolation) Notice and Right to Petition for Habeas Corpus Relief". | • Group 1<br>• Group 2<br>• Group 3<br>• Group 4<br>• Overall |
| To estimate the influence of potential environmental factors in participants who completed the 2-dose primary vaccination series | The number and proportion of participants based on the following potential environmental factors:<br>• The definition of co-residents: Person(s) live(s) at the same address with the participant for more than one day<br>• Number and proportion of participants without COVID-19, living with co-residents who infected withSARS-COV-2.<br>• Number and proportion of participants' infectious period which overlapped the infectious period of co-residents within 14 days | • Group 1<br>• Group 2<br>• Group 3<br>• Group 4<br>• Overall |
| To evaluate influence factors for the time to first confirmed SARS-COV-2 infection after 2- | The time to first confirmed SARS-COV-2 infection after 2-dose primary vaccination series affected by the following factors: | • Group 1<br>• Group 2<br>• Group 3<br>• Group 4 |

| | | |
|---|---|---|
| dose primary vaccination series | • Age<br>• Sex<br>• Comorbidity<br>• Obesity (BMI≥30 kg/$m^2$) | • Overall |
| Compared with external information, incidence rate of COVID-19 | The comparison between the external information described in section 8.4 of protocol and the data collected in this study in terms of the following items:<br>• incidence rate of COVID-19<br>• The rate of breakthrough infections after primary vaccination series<br>• The rate of breakthrough infections after the booster vaccination | • Group 1<br>• Group 2<br>• Group 3<br>• Group 4 |

Definition of study groups is as below.

| Group | Description |
|---|---|
| Group 1 | Participants who received two doses of UB-612 and did not receive COVID-19 vaccines other than UB-612 before study enrolment |
| Group 2 | Participants who received two doses of UB-612 and then received COVID-19 vaccine(s) other than UB-612 before study enrolment |
| Group 3 | Participants who received three doses of UB-612 and did not receive COVID-19 vaccines other than UB-612 before study enrolment |
| Group 4 | Participants who received three doses of UB-612 and then received COVID-19 vaccine(s) other than UB-612 before study enrolment |
| Overall | Pooling Group 1 to Group 4 |

### 7. INVESTIGATIONAL PLAN

#### 7.1. Study Design – General Aspects

This is a questionnaire to survey the COVID-19 breakthrough infection in adult and adolescent volunteers completed V-205 phase II study. Study population consists of younger and elderly adult and adolescent subjects who had ever received at least two UB-612 vaccinations (the 1st dose and the 2nd dose) in the V-205 phase II study. Eligible subjects will be contacted by telephone to obtain the consent. The questionnaire contains those participants' SARS-COV-2 infection history and corresponding disease severity after UB-612 2-dose primary vaccination series.

#### 7.2. Discussion of Study Design

The primary objective of this study is to survey the disease severity of COVID-19 cases post UB-612 primary series. Secondary objectives are to obtain the numbers of COVID-19 cases post UB-612 primary series, to describe risk factors for developing COVID-19 and developing moderate and severe COVID-19 illness, and to investigate the time to first occurrence of COVID-19 after 2-dose primary vaccination series.

In order to understand the COVID-19 breakthrough preliminarily after UB-612 primary vaccination series, e-questionnaire will be used in this study. Using online questionnaires can reduce research cost, save research time and increase data accessibility. In this questionnaire survey, the 1st question was aimed to double-checked whether subjects are willing to participate into this study, the 2nd question reflected whether subjects received other COVID-19 booster vaccines, the 3rd and 4th questions collected the information of COVID-19 breakthrough infection, and the 5th to 7th questions surveyed the information of SARS-COV-2 infection in the co-residents of subjects.

#### 7.3. Selection of Study Population

##### 7.3.1. Number of Planned Subjects

Up to 3654 subjects who previously received at least 2 doses of UB-612 vaccination in V-205 phase II study, including 3300 adults and 354 adolescents, will be recruited into this study.

##### 7.3.2. Inclusion Criteria

To be eligible for study entry, subjects must satisfy all of the following inclusion criteria:

1. Male or female who previously participated in and completed at least 2-dose primary

vaccination (1st and 2nd dose) in the V-205 phase II Study.

2. Participants or participants' legal representative must agree to join the questionnaire survey.

3. Participant or the participant's legal representative must understand and agrees to complete the e-questionnaire.

### 8. STUDY CONDUCT

#### 8.1. Procedures

The subjects will be contacted by telephone to inquire the consent to participate into this study. The eligible subjects will get their access right to the pre-designed e-questionnaire. The COVID-19 breakthrough cases with severity and relative information will be collected.

#### 8.2. Demographics / Other Baseline Characteristics

The demographic and other baseline characteristic data for subjects will be extracted from the database of V-205 phase II study.

- **Demographics:** date of birth, age, sex and ethnicity.
- **Study Vaccine Administration:** date of vaccination
- **Risk factor:** comorbidity (collected from V-205 clinical significant medical history), obesity (BMI ≥ 30 $kg/m^2$)

#### 8.3. E-Questionnaire

The e-questionnaire is a 7-item instrument which collects the personal COVID-19 immunization (except UB-612), breakthrough date after the $2^{nd}$ dose UB-612 vaccination, severity of COVID-19 symptoms, number of co-residents, condition of SARS-COV-2 infection in co-residents. The content of e-questionnaire is listed below:

**Questionnaire**

1. Would you like to complete this questionnaire?
   ☐ Yes ☐ No

2. Except UB-612, did you receive any other COVID-19 vaccine?

   ☐ None
   ☐ Received one dose of other COVID-19 vaccine and please fill in the day of non-UB-612 COVID-19 vaccination; Notice: ________________(yyyy/mm/dd)
   ☐ Received two doses of other COVID-19 vaccine and please fill in the day of non-UB-612 COVID-19 vaccination; Notice: ________________(yyyy/mm/dd)
   ☐ Received three doses of other COVID-19 vaccine and please fill in the day of non-UB-612 COVID-19 vaccination; Notice: ________________(yyyy/mm/dd)
   ☐ Received four doses of other COVID-19 vaccine and please fill in the day of non-UB-612 COVID-19 vaccination; Notice: ________________(yyyy/mm/dd)

3. Have you been infected with SAS-COV-2 and received COVID-19 Designated Residence Isolation (Home Isolation) Notice after receiving the 2nd dose of UB-612 vaccine?
   - ☐ Yes, please fill in the first day of the COVID-19 Designated Residence Isolation (Home Isolation)
     Notice: _______________(yyyy/mm/dd)
   - ☐ No (Go to question 5 and 6，and question 7 will not be shown in e- questionnaire)

4. If yes on question 3, what is the severity of your COVID-19 symptoms:
   - ☐ Asymptomatic infection
   - ☐ Mild: had symptoms without hospitalization
   - ☐ Moderate: hospitalized due to infection
   - ☐ Severe: hospitalized in intensive care unit due to infection

5. Do you have any co-resident?

   The definition of co-residents: Person(s) live(s) at the same address with the participant for more than one day.

   - ☐ Yes
     - ☐ 1 co-resident
     - ☐ 2 co-residents
     - ☐ 3 co-residents
     - ☐ 4 co-residents
     - ☐ Equal to or more than 5 co-residents.
   - ☐ No (complete the questionnaire )
   - ☐ Refuse to answer

6. Has your co-resident ever been infected with SARS-COV-2?

   - ☐ Yes
     - ☐ 1 co-resident was infected with SARS-COV-2
     - ☐ 2 co-residents were infected with SARS-COV-2
     - ☐ 3 co-residents were infected with SARS-COV-2
     - ☐ 4 co-residents were infected with SARS-COV-2
     - ☐ Equal to or more than 5 co-residents infected with SARS-COV-2
   - ☐ No (complete the questionnaire )
   - ☐ Refuse to answer

7. Did you have confirmed SARS-COV-2 infection within 14 days before or after the date upon which any co-residents had confirmed COVID-19?

   ☐ Yes

   ☐ No

   ☐ I'm not sure

   ☐ Refuse to answer

**8.4. External Information about breakthrough infection**

There will be historical breakthrough infection data collected in terms of the following items:

- Incidence rate of COVID-19
- The rate of breakthrough infections after primary vaccination series
- The rate of breakthrough infections after the booster vaccination

Those data will be collected mainly from source data of Taiwan CDC that will be offered and analyzed by UBI Asia, Inc.

### 9. STATISTICAL METHODS

#### 9.1. Statistical and Analytical Plans

A statistical analysis plan will be prepared and finalized prior to database lock of the study. The statistical analysis plan will include full details of all planned statistical analyses.

#### 9.2. General Consideration

All the data summary is based on subjects who have the valid e-questionnaire record. The available data will be displayed via the descriptive statistics no statistical testing is adopted. For continuous variables, the number, mean, standard deviation, median, minimum, and maximum values will be presented. For categorical variables, the numbers and percentages of subjects in each class will be presented. All confidence intervals, if provided, will be the 95% confidence intervals.

#### 9.3. Demographic and Other Baseline Characteristics

Demographic and baseline characteristic data extracted from the database of V-205 phase II study will be summarized. Descriptive statistics (N, mean, standard deviation, median, minimum, and maximum) will be presented for continuous variables. The number and percentage of subjects in each category will be presented for categorical variables. No formal testing of demographic or baseline characteristics will be performed.

#### 9.4. Analysis of Primary Endpoint

- The number and percentage of COVID-19 cases with the occurrence of the severity

  The primary endpoint of COVID-19 cases with the occurrence of the severity will be descriptively summarized with number and percentage of COVID-19 cases in severity by groups defined in section 6. The 95% Clopper-Pearson confidence interval will be also presented.

#### 9.5. Analysis of Secondary Endpoints

- The number of participants with the occurrence of COVID-19 cases confirmed by receiving "COVID-19 Designated Residence Isolation (Home Isolation) Notice and Right to Petition for Habeas Corpus Relief"

  The number and percentage of COVID-19 cases will be summarized along with 95% Clopper-Pearson confidence interval by groups defined in section 6.

- The number and proportion of COVID-19 based on the following risk factors

The number and proportion of COVID-19 based on the risk factors of age, sex, comorbidity, and obesity (BMI≥30 kg/m$^2$) will be summarized along with 95% Clopper-Pearson confidence interval by groups defined in section 6. The risk factor of age will be categorized as adults, younger adults, elderly adults, and adolescents defined in V-205 phase II study. In addition, the risk factor of comorbidity is collected the record of clinical significant medical history as "Any tumor (other than leukemia, lymphoma, metastatic solid tumor)", "Cerebrovascular disease", "Dementia", "Diabetes", "Mild liver disease (inflammation, hepatitis)", "Moderate or severe renal disease (eGFR: 30-59 for moderate, eGFR: <30 for severe)", "Myocardial infarction", "Peripheral vascular disease", and "Ulcer disease" from V-205 phase II study.

- The number and proportion of moderate and severe COVID-19 illness based on the following risk factors

  The number and proportion of moderate and severe COVID-19 illness based on the risk factors of age, sex, comorbidity, and obesity (BMI≥30 kg/m2) will be summarized along with 95% Clopper-Pearson confidence interval by groups defined in section 6.

- The time to first confirmed SARS-COV-2 infection after 2-dose primary vaccination series and before other vaccination

  The time to first confirmed SARS-COV-2 infection after 2-dose primary vaccination series is defined as the time from the date completed 2-dose primary vaccination series to the date of first confirmed SARS-COV-2 infection. The date of first COVID-19 is defined as the 1$^{st}$ day of residence isolation recorded in "COVID-19 Designated Residence Isolation (Home Isolation) Notice and Right to Petition for Habeas Corpus Relief". If a subject has no confirmed SARS-COV-2 infection at the study cut-off date, the subject is censored at the date he or she filled out the study questionnaire.

  The Kaplan-Meier approach will be used to analysis the time to event data with median time and corresponding 95% confidence interval. Draw the Kaplan-Meier plot to demonstrate the progress of time to the first confirmed SARS-COV-2 infection after 2-dose primary vaccination series.

  For subjects who had no COVID-19, the duration after 2-dose primary vaccination series to the E-questionnaire completion will be summarized by descriptive statistics.

##### 9.6. Analysis of Exploratory Endpoints

The outcome measure of exploratory endpoints will be incidence rate of COVID-19 cases, the infected risk with potential environmental factors and time to event period with influence factors (including age, sex, comorbidity and obesity). The differences between subgroups will be analyzed by Student's t test or chi-square test.

- The incidence rate of participants presented as a percentage with the occurrence of COVID-19 cases

  The incidence rate of participants presented as a percentage with the occurrence of COVID-19 cases will confirm by who receiving "COVID-19 Designated Residence Isolation (Home Isolation) Notice and Right to Petition for Habeas Corpus Relief". New cases of COVID-19 occurred in the total subject population between the periods after the 2-dose primary vaccination series to the E-questionnaire completion. The incidence rate of COVID-19 cases the standard deviation, median, minimum, and maximum values will be presented.

- The number and proportion of participants based on the potential environmental factors

  The potential environmental factors will evaluate via co-resident infection. The co-residents will define as person(s) live(s) at the same address with the participant for more than one day. Number and proportion of participants without COVID-19, living with co-residents who infected with COVID-19 will be summarized along with 95% Clopper-Pearson confidence interval by groups defined in section 6.

  Number and proportion of participants' infectious period which overlapped the infectious period of co-residents within 14 days will be summarized along with 95% Clopper-Pearson confidence interval by groups defined in section 6.

- The time to first confirmed SARS-CoV-2 infection after 2-dose primary vaccination series affected by influence factors

  This object will be investigating the relationship of uninfected period and influence factors. For subject who had COVID-19, the uninfected period will base on the date till receiving "COVID-19 Designated Residence Isolation (Home Isolation) Notice and Right to Petition for Habeas Corpus Relief". For subjects who had no COVID-19, the duration after 2-dose primary vaccination series to the E-questionnaire completion. The number and proportion of COVID-19 based on the risk factors of age, sex, comorbidity, and obesity (BMI$\geq$30 kg/m$^2$) will be summarized along with 95% Clopper-Pearson confidence interval by groups defined in section 6.

- The comparison between the external information

  The comparison between the external information described in section 8.4. The prevalence rate and breakthrough infections of COVID-19 in Taiwan will be applied as reference.

##### 9.7. Handling of Missing Data

No imputation will be considered for the missing observations from the subjects reported e-questionnaire.

### 10. QUALITY ASSURANCE AND QUALITY CONTROL

#### 10.1. Audit and Inspection

Study centers and study documentation may be subject to Quality Assurance audit during the course of the study by the sponsor or its nominated representative. In addition, inspections may be conducted by regulatory authorities at their discretion.

Data for each subject will be recorded on an e-questionnaire. This study might qualify for a waiver of informed consent under the permission of site's Institutional Review Board (IRB), or Ethics Committee (EC). However, if the site's IRB sees a written informed consent is required, an informed consent form will be provided.

Due to study data collected directly by e-questionnaire, the study monitor activity may be omitted unless necessary. However, all study procedures will be under the current good clinical practice (cGCP) and International Council for Harmonisation (ICH) guidelines to ensure that the data collected in e-questionnaire are accurate and reliable.

The investigator must permit the IEC/IRB, the sponsor's internal auditors, and representatives from regulatory authorities directly access to all study-related documents for confirmation of data contained within the e-questionnaire.

#### 10.2. Data Management and Coding

The sponsor and/or the appointed representative(s) will be responsible for activities associated with the data management of this study. This will include setting up a relevant database and data transfer mechanisms, along with appropriate validation of data and resolution of queries. Data generated within this clinical study will be handled according to the relevant standard operating procedures of the data management and biostatistics departments of the sponsor and/or the appointed representative(s).

Study centers will complete the eCRF. Data entered into the eCRF must be verifiable against source documents at the study center. Data to be recorded directly on the eCRF will be identified and the eCRF will be considered the source document. Any changes to the data entered into the data capture system will be compliant to FDA CFR 21 Part 11.

Missing or inconsistent data will be queried to the investigator for clarification. Subsequent modifications to the database will be documented.

#### 11. RECORDS AND SUPPLIES

##### 11.1. Drug Accountability

Not applicable.

##### 11.2. Financing and Insurance

Financing of this study will be outlined in a separate agreement between the contract research organization and the sponsor. Insurance will be omitted in this study.

### 12. ETHICS

#### 12.1. Independent Ethics Committee or Institutional Review Board

Before initiation of the study at each study centre, the protocol, including questionnaire in Chinese, will be submitted to the appropriate IEC/IRB. Written approval of the study and all relevant study information must be obtained before the study centre can be initiated. Any necessary extensions or renewals of IEC/IRB approval must be obtained for changes to the study such as amendments to the protocol, or other study documentation. The written approval of the IEC/IRB together must be filed in the study files.

The investigator will submit written summaries of the study status to the IEC/IRB as required. On completion of the study, the IEC/IRB will be notified that the study has ended.

#### 12.2. Regulatory Authorities

Relevant study documentation will be submitted to the regulatory authorities under request.

#### 12.3. Ethical Conduct of the Study

The investigator(s) and all parties involved in this study should conduct the study in adherence to the ethical principles based on the Declaration of Helsinki, cGCP, ICH guidelines, and the applicable national and local laws and regulatory requirements.

#### 12.4. Informed Consent

Due to this study is non-intervention and retrospective, this study might be qualified for a waiver of written informed consent under the permission of site's IEC(s)/IRB(s). Under this process, subjects will be asked for consent in telephone by the investigator or designated personnel, and double-check their willingness in e-questionnaire.

However, if the site's IRB sees a written informed consent is required, an informed consent form will be provided.

The investigator is responsible for ensuring that no subject undergoes any study related examination or activity before that subject has given informed consent to participate in the study.

The investigator or designated personnel will inform the subject of the objectives, methods, anticipated benefits and potential risks and inconveniences of the study. The subject should be given every opportunity to ask for clarification of any points s/he does not understand and, if necessary, ask for more information. At the end of the interview, the subject will be given ample time to consider the study.

It should be emphasized that the subject may refuse to enter the study or to withdraw from the study at any time, without consequences for their further care or penalty or loss of benefits to which the subject is otherwise entitled. Subjects who refuse to give or who withdraw informed consent should not be included or continue in the study.

If new information becomes available that may be relevant to the subject's willingness to continue participation in the study, those will be approved by the IEC(s)/IRB(s) (and regulatory authorities, if required). The study subjects will be informed about this new information and reconsent will be obtained.

##### 12.5. Subject Confidentiality

Auditors, and other authorized agents of the sponsor and/or its designee, the IEC(s)/IRB(s) approving this research, and the United States (US) FDA, as well as that of any other applicable agency(ies), will be granted direct access to the study subjects' questionnaire data, without violating the confidentiality of the subjects to the extent permitted by the law and regulations. In any presentations of the results of this study or in publications, the subjects' identity will remain confidential.

All personal data collected and processed for the purposes of this study should be managed by the investigator and his/her staff with adequate precautions to ensure confidentiality of those data, applicable to national and/or local laws and regulations on personal data protection.

#### 13. REPORTING AND PUBLICATION, INCLUDING ARCHIVING

After completion of the study (end of study defined as the date of the last visit of the last subject), all documents and data relating to the study will be kept in an orderly manner by the investigator in a secure study file. This file will be available for inspection by the sponsor or its representatives. Essential documents should be retained for 2 years after the final marketing approval in an ICH region or for at least 2 years since the discontinuation of clinical development of the investigational product. It is the responsibility of the sponsor to inform the study centre when these documents no longer need to be retained. The investigator must contact the sponsor before destroying any study related documentation. In addition, all subject medical records and other source documentation will be kept for the maximum time permitted by the hospital, institution, or medical practice.

The sponsor must review and approve any results of the study or abstracts for professional meetings prepared by the investigator(s). Published data must not compromise the objectives of the study. Data from individual study centres in multicenter studies must not be published separately.

**Appendix 2. IRB Approval Tracker: Ethical Committee Endorsements for the Questionnaire-based, Retrospective, Phase-2 Extension Study**

| **Protocol title** | A Questionnaire-based, Retrospective, Phase-2 Extension Study to Investigate the COVID-19 Breakthrough Infection after 2 doses of UB-612 Vaccination in Adolescent, Younger and Elderly Adult Volunteers | |
|---|---|---|
| Ethics Committee / Institutional Review Board (IRB) | Decision made by ethics oversight body | Approved Date |
| Changhua Christian Hospital | Approved | 2022-12-27 |
| Chang Gung Medical Foundation | Approved | 2023-01-19 |
| China Medical University Hospital | Approved | 2022-11-08 |
| Far Eastern Memorial Hospital | Approved | 2022-12-19 |
| Kaohsiung Medical University Chung-Ho Memorial Hospital | Approved | 2022-12-22 |
| National Cheng Kung University Hospital | Approved | 2023-01-01 |
| Taipei Medical University | Approved | 2022-12-22 |
| Tri-Service General Hospital | Approved | 2022-12-22 |
| Kaohsiung Veterans General Hospital | Approved | 2022-12-18 |
| Taichung Veterans General Hospital | Approved | 2023-01-09 |
| Taipei Veterans General Hospital | Approved | 2022-12-22 |
